# Mental health in a diverse sample of healthcare workers during the COVID-19 pandemic: cross-sectional analysis of the UK-REACH study

**DOI:** 10.1101/2022.02.03.22270306

**Authors:** Carl A Melbourne, Anna L Guyatt, Laura Nellums, Padmasayee Papineni, Amit Gupta, Irtiza Qureshi, Christopher A Martin, Luke Bryant, Catherine John, Mayuri Gogoi, Fatimah Wobi, Amani Al-Oraibi, Jonathan Chaloner, Avinash Aujayeb, Bindu Gregary, Susie Lagrata, Rubina Reza, Sandra Simpson, Stephen Zingwe, Martin Tobin, Sue Carr, Kamlesh Khunti, Laura J Gray, I Chris McManus, Katherine Woolf, Manish Pareek, the UK-REACH collaborative group

**Author notes:** indicates equal contribution (Carl A Melbourne, Anna L Guyatt). Manish Pareek (Chief investigator), Laura J Gray (University of Leicester), Laura Nellums (University of Nottingham), Anna L Guyatt (University of Leicester), Catherine John (University of Leicester), I Chris McManus (University College London), Katherine Woolf (University College London), Ibrahim Akubakar (University College London), Amit Gupta (Oxford University Hospitals), Keith R Abrams (University of York), Martin D Tobin (University of Leicester), Louise V Wain (University of Leicester), Sue Carr (University Hospital Leicester), Edward Dove (University of Edinburgh), Kamlesh Khunti (University of Leicester), David Ford (University of Swansea), Robert Free (University of Leicester). Author contributions Conception or design of the work: LB, SC, AG, ALG, LJG, CJ, KK, CAM, CM, ICM, LN, PP, MP, IQ, MDT, KW. Acquisition of the data: AA, LB, SC, ALG, BG, CJ, CM, ICM, MP, RR, SS, KW, SZ. Analysis of the data: ALG, CM. Interpretation of data for the work: AG, ALG, CM, LN, PP, MP, IQ, KW. Drafting the work: AG, ALG, CM, LN, PP, MP, IQ, KW. Revising it critically for important intellectual content: SC, AG, ALG, LJG, CJ, KK, CAM, CM, ICM, LN, PP, MP, IQ, MDT, KW. Final approval of the version to be published; agreement to be accountable for all aspects of the work in ensuring that questions related to the accuracy or integrity of any part of the work are appropriately investigated and resolved: All authors. Guarantors: CM, ALG, MP. The guarantors accept full responsibility for the work and/or the conduct of the study, had access to the data, and controlled the decision to publish. The corresponding author attests that all listed authors meet authorship criteria and that no others meeting the criteria have been omitted. The lead authors affirm that the manuscript is an honest, accurate, and transparent account of the study being reported; that no important aspects of the study have been omitted; and that any discrepancies from the study as originally planned have been explained. Role of the funding source The funding source was not involved in study design, data collection or analysis, interpretation, or writing of the report. Data sharing Deidentified participant data are available via a system of managed access, to access data or samples produced by the UK-REACH study please contact to discuss your request. The protocol for the UK-REACH longitudinal cohort study is available at: http://dx.doi.org/10.1136/bmjopen-2021-050647, and a Data Dictionary of the baseline questionnaire data is available at: https://uk-reach.org/main/data-dictionary/. Competing interests SC is Deputy Medical Director of the General Medical Council, UK Honorary Professor, University of Leicester. MDT receives funding from GSK for collaborative research projects outside of the submitted work. KK is Director of the University of Leicester Centre for Black Minority Ethnic Health, Trustee of the South Asian Health Foundation and Chair of the Ethnicity Subgroup of the UK Government Scientific Advisory Group for Emergencies (SAGE). MP reports grants from Sanofi, grants and personal fees from Gilead Sciences and personal fees from QIAGEN, outside the submitted work.

## Abstract

**Objectives:** To investigate how ethnicity and other sociodemographic, work, and physical health factors are related to mental health in UK healthcare and ancillary workers (HCWs), and how structural inequities in these factors may contribute to differences in mental health by ethnicity.

**Design:** Cross-sectional analysis of baseline data from the UK-REACH national cohort study

**Setting:** HCWs across UK healthcare settings.

**Participants:** 11,695 HCWs working between December 2020-March 2021.

**Main outcome measures:** Anxiety or depression symptoms (4-item Patient Health Questionnaire, cut-off ≥3), and Post-Traumatic Stress Disorder (PTSD) symptoms (3-item civilian PTSD Checklist, cut-off >5).

**Results:** Asian, Black, Mixed/multiple and Other ethnic groups had greater odds of PTSD than the White ethnic group. Differences in anxiety/depression were less pronounced. Younger, female HCWs, and those who were not doctors had increased odds of symptoms of both PTSD and anxiety/depression. Ethnic minority HCWs were more likely to experience the following work factors that were also associated with mental ill-health: workplace discrimination, feeling insecure in raising workplace concerns, seeing more patients with COVID-19, reporting lack of access to personal protective equipment (PPE), and working longer hours and night shifts. Ethnic minority HCWs were also more likely to live in a deprived area and have experienced bereavement due to COVID-19. After adjusting for sociodemographic and work factors, ethnic differences in PTSD were less pronounced and ethnic minority HCWs had lower odds of anxiety/depression compared to White HCWs.

**Conclusions:** Ethnic minority HCWs were more likely to experience PTSD and disproportionately experienced work and sociodemographic factors associated with PTSD, anxiety and depression. These findings could help inform future work to develop workplace strategies to safeguard HCWs’ mental health. This will only be possible with adequate investment in staff recruitment and retention, alongside concerted efforts to address inequities due to structural discrimination.

**Summary box:** *What is already known on this topic:*

- The pandemic is placing healthcare workers under immense pressure, and there is currently a mental health crisis amongst NHS staff
- Ethnic inequities in health outcomes are driven by structural discrimination, which occurs inside and outside the workplace
- Investigating ethnic inequities in the mental health of healthcare workers requires large diverse studies, of which few exist

*What this study adds:*

- In UK-REACH (N=11,695), ethnic minority staff had higher odds of Post-Traumatic Stress Disorder symptoms; we report many other factors associated with mental-ill health, including those experienced disproportionately by ethnic minority staff, such as workplace discrimination, contact with more patients with COVID-19, and bereavement due to COVID-19
- These findings underline the moral and practical need to care for staff mental health and wellbeing, which includes tackling structural inequities in the workplace; improving staff mental health may also reduce workforce understaffing due to absence and attrition

## Introduction

The COVID-19 pandemic is a time of immense pressure for healthcare workers (HCWs) in both clinical and ancillary roles.

In early 2020, the threat of the impending pandemic escalated rapidly into many critically unwell patients in hospitals,^1^ major increases in demand in the community,^2^ longer working hours, constantly evolving restrictions, and changes to working practices and services.^3^ These pressures are alongside the risk of HCWs’ own COVID-19 illness or death in addition to that of their patients, colleagues and family and friends, and the possibility of infecting others, compounded by shortages in personal protective equipment (PPE)^4^. HCWs have found their personal moral codes threatened or violated, by being constrained in their ability to take action(s) that they consider ethically correct, for example due to resource pressures, or institutional culture.^5 6^ Many of these complex factors have been shown to be associated with higher rates of anxiety, depression and post-traumatic stress disorders among HCWs both in the UK and worldwide during the pandemic.^7 8^

The work stressors borne by HCWs are in addition to those experienced at the population level,^9^ such as caring responsibilities, financial difficulties, pre-existing health needs (which may have led vulnerable HCWs to not be able to work, or to work remotely^10^), and reduction in social support due to lockdown restrictions.

Ethnic minority communities and HCWs have experienced higher rates of infection and mortality from COVID-19.^11–14^ In addition, many NHS healthcare workers from ethnic minority groups have family and friends overseas who will have been impacted by pandemic waves globally. Adjusting statistically for geographical, socio-demographic, and pre-pandemic health factors substantially reduces differences in mortality between most ethnic minority groups and the White population, which may suggest that structural factors partly mediate such differences.^13 15^ In the NHS and broader society, structural inequities by ethnicity are prevalent.^16 17^ The 2020 NHS staff survey reported that more ethnic minority HCWs than White HCWs experienced workplace harassment, bullying or abuse in 2020, and the gap widened from 2017-2020^18^. The relationship between these experiences and poor mental health are well-known.^19^ Indirect measures of inequity at work, e.g. both quantitative (workload) and qualitative (roles at work, trust in employer), are less studied, but are described as work predictors for common mental health problems, alongside external factors.^20 21^ We therefore hypothesised that ethnic inequities in stressful conditions at work and factors outside of work could contribute to differences in mental health between ethnic groups.

We used baseline questionnaire data from the United Kingdom Research study into Ethnicity and COVID-19 outcomes in Healthcare workers (UK-REACH) to investigate three research questions: i) what are the associations between ethnicity, other sociodemographic, work and physical health factors and symptoms of anxiety/depression and Post-Traumatic Stress Disorder (PTSD)? ii) what evidence is there of ethnic inequities in these factors? and iii) how does adjusting for these factors alter the association between ethnicity and mental health?

## Methods

### Study population

UK-REACH includes six work streams that aim to examine the impact of the COVID-19 pandemic on HCWs, and to explore potential disparities by ethnicity.

The current cross-sectional study uses baseline questionnaire data from participants in the UK-REACH prospective cohort study. Details of the cohort study design and participant recruitment are given in the study protocol,^22^ and details of the questionnaire are available at https://www.uk-reach.org/data-dictionary.

Briefly, clinical and ancillary HCWs aged 16 or over, living in the four nations of the UK, and from diverse ethnic backgrounds were invited to participate, either by one of seven healthcare professional regulators, a participating NHS trust or by broader study advertisement. Participants who gave their informed consent to join the study were invited to complete the baseline questionnaire online (data collected between 4^th^ December 2020 - 8^th^ March 2021).

### Outcome measures

The primary outcomes were symptoms of anxiety or depression (anxiety/depression) combined due to their comorbid nature, and PTSD, as measured by screening tools. Anxiety and depression symptoms were measured using the Generalised Anxiety Disorder 2-item scale (GAD-2) and the Patient Health Questionnaire-2 scale (PHQ-2), respectively. Scores were combined (as the PHQ-4), with a score ≥3 used to derive a pooled outcome of potential anxiety or depression.^23^ Symptoms of PTSD were evaluated using a three-item version of the PTSD Checklist—civilian version (PCL-C), with a score ≥5 used to define a dichotomised outcome.^24^

### Predictor variables

We studied multiple factors related to participants’ lives, both at work and outside of work, for their association with either of the mental health outcomes. These variables were chosen *a priori*, because of published or hypothesised associations with worse mental health. For a full list and details of how individual variables were measured and coded, see **Supplementary Text**.

Key demographic variables studied were ethnicity, age and sex. Self-reported ethnicity was categorised into five broad groups to maximise power for statistical analysis, consistent with categorisation used for the 2011 UK Census^25^ (White, Mixed/multiple ethnic groups, Asian/Asian British, Black/African/Caribbean/Black British, Other ethnic group, hereafter abbreviated to ‘White’ ‘Mixed/multiple’, ‘Asian’, ‘Black’ and ‘Other’). In this paper, we use the term ethnic minorities to describe participants belonging to the ‘Mixed/multiple’, ‘Asian’, ‘Black’ and ‘Other’ groups.

Other socioeconomic factors studied were: index of multiple deprivation (IMD) decile,^26^ being born in the UK, the number of other individuals in the participant’s household, whether participants live with children, whether participants live with adults ≥65 years, and bereavement due to COVID-19.

Physical health factors studied were: number of physical long-term conditions, SARS-CoV-2 infection status, frequency of alcohol consumption, smoking status, and physical activity (measured by the GP Physical Activity Index).^27^

Working factors studied were: job role (doctor or medical support; nurse, nursing associate or midwife; allied health professional; dental; admin, estates or other), type of workplace (e.g. intensive care unit), COVID-19 redeployment status, whether or not a participant had experienced discrimination at work, whether a participant would feel secure to raise concerns at work about unsafe clinical practice, and whether a participant would trust their organisation to address their concerns. We also included variables measuring day-to-day characteristics of participants’ job role, including degree of patient contact per week (numbers of patients with COVID-19, and number without COVID-19), weekly working hours, frequency of night working, and access to appropriate PPE.

## Statistical analysis

Given the focus of the UK-REACH study on understanding disparities in outcomes by ethnicity, only individuals with complete data for ethnicity and at least one of the mental health outcome measures of anxiety/depression (GAD-2, PHQ-2) or PTSD (PCL-C) scales were retained for this analysis. For the main analysis, the sample was further restricted to those HCWs who reported that they were working at the time of baseline questionnaire completion, since the aim was to study the association between working factors and mental health.

To account for missing data for the remaining variables, we performed multiple imputation and used Rubin’s rules^28^ to combine estimates and standard errors from ten imputations to produce a final set of results. Due to lack of comparability of IMD across countries,^26^ we opted to impute an English IMD entry for those outside of England rather than use Welsh, Scottish and Northern Irish IMD values.

We report descriptive statistics for predictor variables as median and IQR for continuous variables, and as percentages for categorical variables.

### Association of sociodemographic, physical health and work factors with anxiety/depression and PTSD

We used logistic regression to calculate associations between ethnicity and other sociodemographic factors, physical health and work factors with each dichotomised mental health outcome. We present both univariable associations, and associations adjusted for age, sex, ethnicity and job role.

### Associations of ethnicity with sociodemographic, physical health and work factors

To explore evidence of inequities in sociodemographic, physical health and work factors by ethnicity, we performed regression analyses, also adjusted for age, sex and job role, to estimate the associations between ethnicity and work/non-work factors (using linear, logistic and ordinal logistic regression for continuous, binary, and ordered categorical outcomes respectively).

### Serial adjustment of association between ethnicity and mental health

We further adjusted associations between ethnicity and mental health outcomes for factors in and outside of work, to explore whether the association attenuated when adjusting for possible mediators. After initially adjusting for variables hypothesised as confounders (model 1), we further adjusted for social (model 2), health (model 3) and work factors (model 4).

### Sensitivity Analyses

To assess the effect of only including individuals who were working at time of questionnaire completion, we recalculated the associations between non-work factors and mental health after additionally including individuals who were not working.

We included “date of consent” (month/year) in the model as an unordered categorical variable, as a proxy for date of questionnaire completion, to explore its effect on original estimates in the main analysis sample, and its association with mental health outcomes.

Since multiple imputation can introduce bias when the reason for why data are missing is unclear, we re-calculated associations between work/non-work factors and mental health after restricting to complete data for confounders for each exposure. We also re-calculated the serially adjusted analysis for associations between ethnicity and mental health, restricting to individuals with complete data for *all* covariates.

### Reporting of results

We do not report results corrected for multiple testing, since this approach emphasises the inappropriate dichotomisation of p-values by arbitrary thresholds. This would also be over-conservative in our analysis due to correlation between variables. Instead, we report the overall pattern of results, reporting exact p-values, and use estimates of effect size and 95% confidence intervals to assess the strength of associations, as recommended previously.^29,30,31^

All analyses were undertaken in Stata 16.^32^

### Ethical approval and engagement

The study was approved by the Health Research Authority (Brighton and Sussex Research Ethics Committee; ethics reference: 20/HRA/4718). All participants gave their informed consent via an online consent process.

### Patient and Public Involvement

UK-REACH includes a Professional Expert Panel of HCWs from a range of ethnic backgrounds, occupations, and genders, as well as a Stakeholder Advisory Group with representatives from national and local organisations (see study protocol).^22^

## Results

### Description of analysed cohort

Of 17,891 individuals recruited to UK-REACH, 15,119 individuals responded to the baseline questionnaire, of whom 12,282 had complete data for ethnicity and the mental health outcomes. Of these, 11,695 were working at the time of baseline questionnaire completion, and these individuals formed the main analysis sample (see **Supplementary Figure 1** for a flowchart describing the sample selection, from recruitment onwards).

**Table 1** presents descriptive statistics for the main analysis sample. Around three-quarters were female, with a median age of 45 years (IQR 34-54). Approximately 30% were from ethnic minority groups (19.1%, 4.2%, 4.2% and 2.1% from Asian, Black, Mixed/multiple and Other ethnic groups, respectively). In terms of professional role, 42% of the cohort were from allied health professions, including pharmacists, 23% were doctors or in medical support roles; 20% were nurses, nursing associates or midwives; 6% were dentists or in other dental roles; 5% were in admin, estates or other roles; and 4% had a missing job role.

**Table 1.**
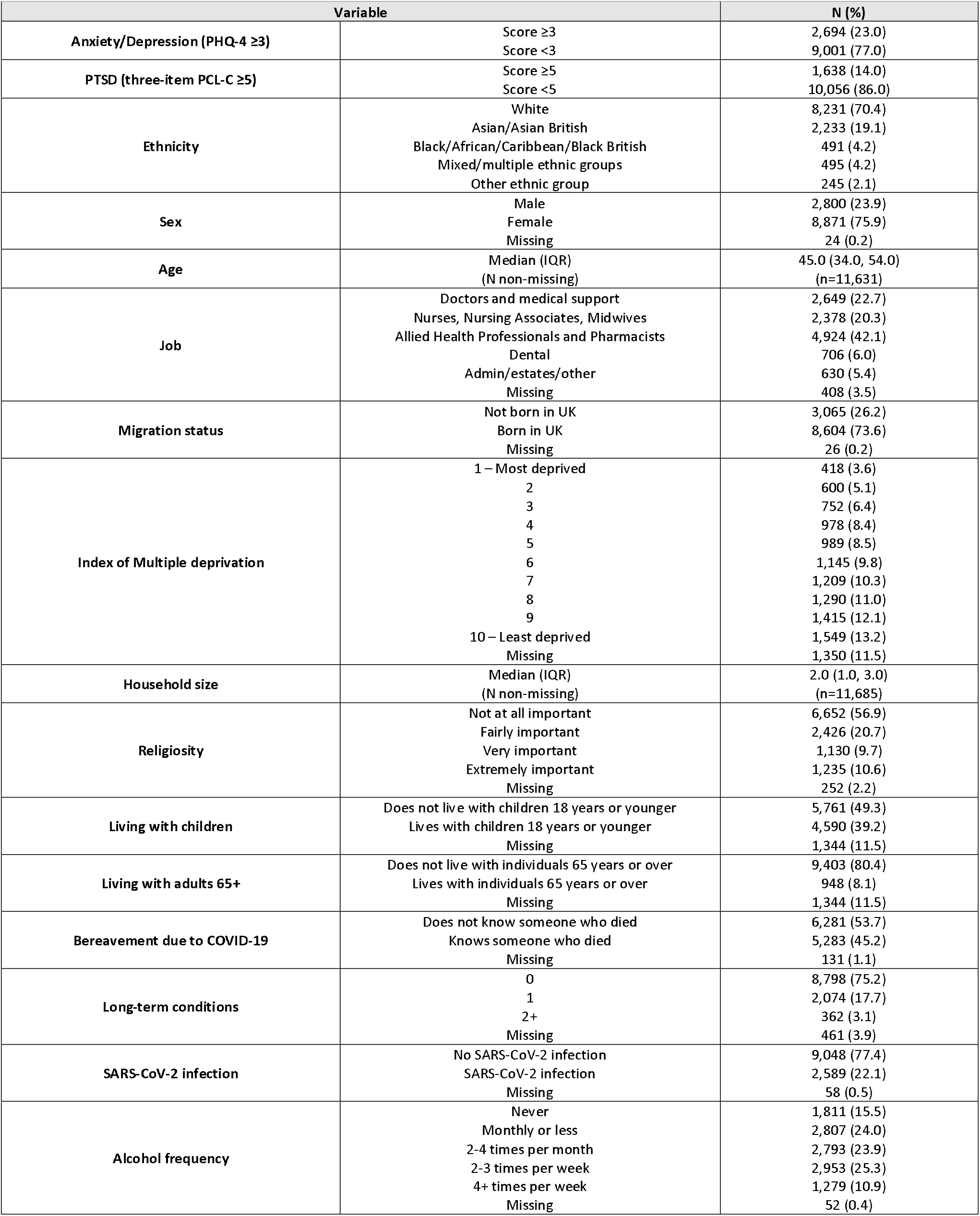
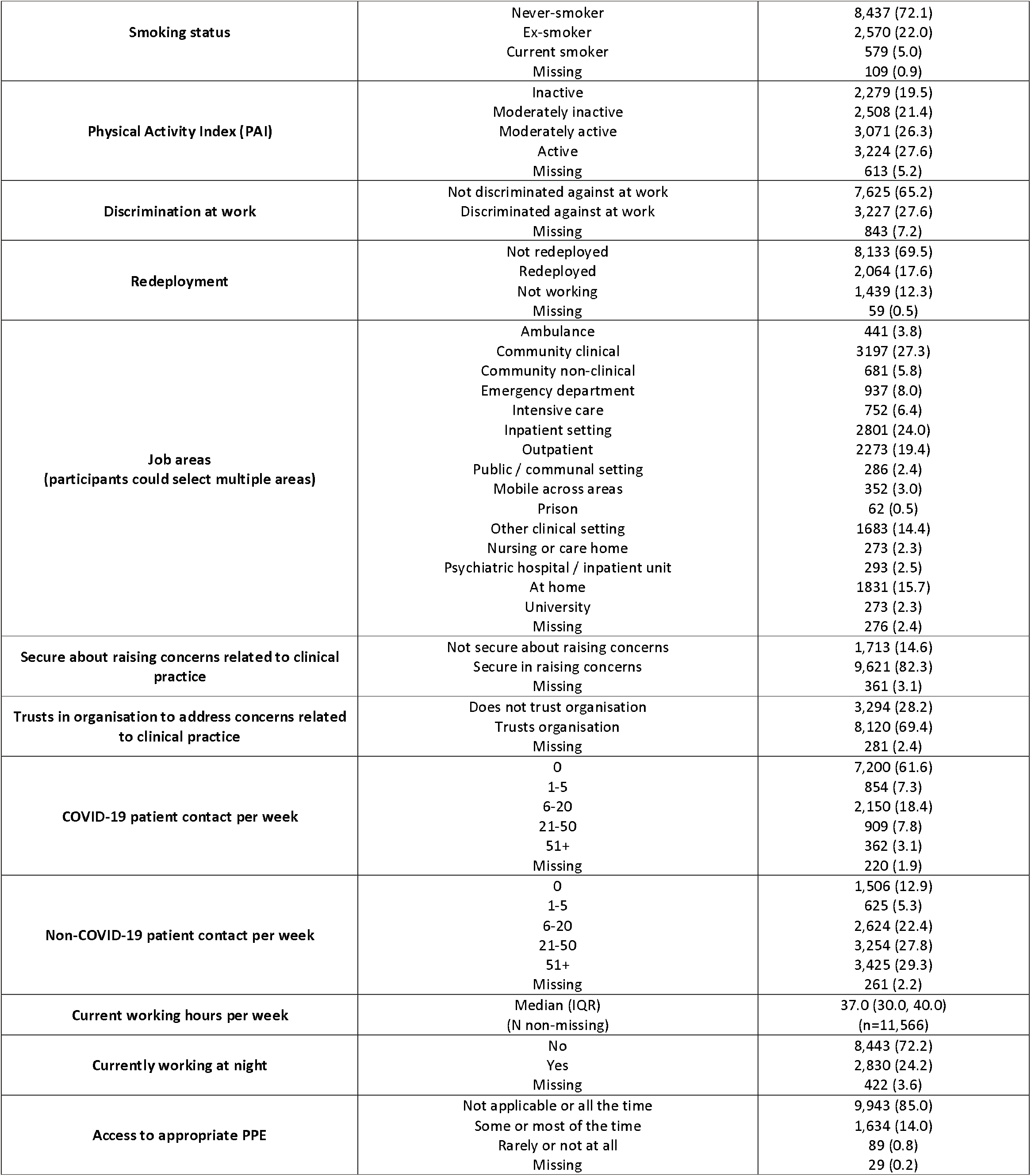
Descriptive of variables and risk factors for main analysis sample of 11,695 individuals.

### Anxiety/depression and PTSD symptoms, and associations with sociodemographic, physical health and work variables

Approximately 23% (N=2,694) of the sample met the screening criteria for possible anxiety/depression, and 14% (N=1,638) for possible PTSD, and 10% (N=1,124) had evidence of both. **Table 2** shows univariable and multivariable logistic regression results for the associations between ethnicity and other sociodemographic factors, physical health and work factors with both anxiety/depression and PTSD symptoms. Multivariable results are adjusted for age, sex, ethnicity and job role.

**Table 2.** Univariable and adjusted associations (adjusted for age, sex, ethnicity and job role) for Anxiety/Depression and PTSD symptom measures.

| Variable | Anxiety/Depression symptoms (PHQ-4 ≥3) |  |  |  | PTSD symptoms (PCL-C ≥5) |  |  |  |
| --- | --- | --- | --- | --- | --- | --- | --- | --- |
|  | Univariable |  | Adjusted<br>(Age, Sex, Ethnicity, Job role) |  | Univariable |  | Adjusted<br>(Age, Sex, Ethnicity, Job role) |  |
|  | OR (95% CI) | P | OR (95% CI) | P | OR (95% CI) | P | OR (95% CI) | P |
| **Ethnicity** |  |  |  |  |  |  |  |  |
| White | ref |  | ref |  | ref |  | ref |  |
| Asian | 0.85 (0.76,0.95) | 0.005 | 0.94 (0.83,1.07) | 0.351 | 1.21 (1.07,1.38) | 0.004 | 1.55 (1.34,1.78) | 2.13E-09 |
| Black | 0.76 (0.60,0.96) | 0.019 | 0.82 (0.65,1.04) | 0.095 | 1.13 (0.87,1.46) | 0.355 | 1.32 (1.01,1.71) | 0.039 |
| Mixed | 0.97 (0.78,1.20) | 0.787 | 0.95 (0.76,1.18) | 0.618 | 1.03 (0.79,1.34) | 0.831 | 1.11 (0.85,1.46) | 0.431 |
| Other | 1.09 (0.81,1.46) | 0.577 | 1.33 (0.98,1.80) | 0.070 | 1.34 (0.95,1.88) | 0.092 | 1.83 (1.29,2.60) | 0.001 |
| **Sex** |  |  |  |  |  |  |  |  |
| Male | ref |  | ref |  | ref |  | ref |  |
| Female | 1.59 (1.43,1.77) | <1E-10 | 1.3 (1.16,1.46) | 8.6E-06 | 1.63 (1.43,1.87) | <1E-10 | 1.41 (1.22,1.63) | 3.2E-06 |
| **Age (per decade)** | 0.69 (0.67,0.72) | <1E-10 | 0.68 (0.66,0.71) | <1E-10 | 0.79 (0.76,0.83) | <1E-10 | 0.79 (0.75,0.83) | <1E-10 |
| **Job** |  |  |  |  |  |  |  |  |
| Doctors and medical support | ref |  | ref |  | ref |  | ref |  |
| Nurses, NAs, Midwives | 1.84 (1.61,2.11) | <1E-10 | 1.87 (1.61,2.18) | <1E-10 | 2.14 (1.82,2.52) | <1E-10 | 2.51 (2.09,3.01) | <1E-10 |
| Allied Health Professionals and pharmacists | 1.47 (1.30,1.66) | <1E-10 | 1.31 (1.14,1.49) | 7.6E-05 | 1.43 (1.23,1.67) | <1E-10 | 1.52 (1.29,1.79) | 5.9E-07 |
| Dental | 1.65 (1.36,2.01) | <1E-10 | 1.46 (1.20,1.79) | 0.0002 | 1.77 (1.39,2.25) | <1E-10 | 1.78 (1.40,2.27) | 3.5E-06 |
| Admin/estates/other | 1.47 (1.19,1.81) | 0.00031 | 1.36 (1.10,1.70) | 0.005 | 1.52 (1.17,1.96) | 0.002 | 1.7 (1.30,2.21) | 0.0001 |
| **Migration status** |  |  |  |  |  |  |  |  |
| Not born in UK | ref |  | ref |  | ref |  | ref |  |
| Born in UK | 1.1 (1.00,1.21) | 0.061 | 0.96 (0.85,1.08) | 0.482 | 0.9 (0.80,1.01) | 0.069 | 0.9 (0.78,1.03) | 0.133 |
| **Index of Multiple Deprivation** | 0.93 (0.91,0.95) | <1E-10 | 0.96 (0.94,0.98) | 3.8E-05 | 0.92 (0.90,0.94) | <1E-10 | 0.94 (0.92,0.96) | 5.3E-07 |
| **Household size** | 0.93 (0.90,0.96) | 0.00001 | 0.93 (0.90,0.96) | 2.1E-05 | 0.96 (0.92,1.00) | 0.056 | 0.95 (0.91,0.99) | 0.015 |
| **Religiosity** |  |  |  |  |  |  |  |  |
| Not at all important | ref |  | ref |  | ref |  | ref |  |
| Fairly important | 0.88 (0.79,0.99) | 0.028 | 0.99 (0.88,1.11) | 0.872 | 1.19 (1.04,1.35) | 0.012 | 1.21 (1.05,1.39) | 0.007 |
| Very important | 0.93 (0.80,1.08) | 0.329 | 1.1 (0.94,1.30) | 0.229 | 1.35 (1.14,1.60) | 0.001 | 1.39 (1.16,1.67) | 0.0003 |
| Extremely important | 0.81 (0.70,0.94) | 0.005 | 0.91 (0.77,1.07) | 0.246 | 1.23 (1.03,1.46) | 0.019 | 1.2 (0.99,1.44) | 0.061 |
| **Living with children** |  |  |  |  |  |  |  |  |
| Does not live with children | ref |  | ref |  | ref |  | ref |  |
| Lives with children | 0.89 (0.81,0.97) | 0.008 | 0.89 (0.81,0.98) | 0.014 | 0.98 (0.88,1.09) | 0.647 | 0.97 (0.87,1.09) | 0.646 |
| **Living with adults 65+** |  |  |  |  |  |  |  |  |
| Does not live with adults 65+ | ref |  | ref |  | ref |  | ref |  |
| Lives with adults 65+ | 0.64 (0.54,0.77) | <1E-10 | 0.87 (0.73,1.05) | 0.153 | 0.85 (0.69,1.05) | 0.123 | 0.98 (0.79,1.22) | 0.847 |

|  | Anxiety/Depression symptoms (PHQ-4 ≥3) |  |  |  | PTSD symptoms (PCL-C ≥5) |  |  |  |
| --- | --- | --- | --- | --- | --- | --- | --- | --- |
|  | Univariable |  | Adjusted<br>(Age, Sex, Ethnicity, Job role) |  | Univariable |  | Adjusted<br>(Age, Sex, Ethnicity, Job role) |  |
| **Bereavement due to COVID-19** |  |  |  |  |  |  |  |  |
| Does not know someone who died | ref |  | ref |  | ref |  | ref |  |
| Knows someone who died | 1.18 (1.08,1.29) | 0.0001 | 1.29 (1.18, 1.42) | 2.6E-08 | 1.48 (1.33,1.64) | <1E-10 | 1.49 (1.34,1.67) | <1E-10 |
| **Long-term conditions** | 1.16 (1.07,1.26) | 0.0004 | 1.31 (1.20, 1.42) | 2.2E-09 | 1.25 (1.13,1.38) | <1E-10 | 1.34 (1.21,1.48) | 2.6E-08 |
| **SARS-CoV-2 infection** |  |  |  |  |  |  |  |  |
| No SARS-CoV-2 infection | ref |  | ref |  | ref |  | ref |  |
| SARS-CoV-2 infection | 1.06 (0.95,1.17) | 0.306 | 0.97 (0.87,1.07) | 0.518 | 1.06 (0.93,1.20) | 0.390 | 0.99 (0.87,1.12) | 0.821 |
| **Alcohol frequency** |  |  |  |  |  |  |  |  |
| Never | ref |  | ref |  | ref |  | ref |  |
| Monthly or less | 1.09 (0.95,1.25) | 0.210 | 0.95 (0.82,1.10) | 0.489 | 0.99 (0.85,1.16) | 0.917 | 0.96 (0.81,1.13) | 0.601 |
| 2-4 times per month | 0.94 (0.82,1.08) | 0.395 | 0.81 (0.69,0.94) | 0.005 | 0.77 (0.65,0.91) | 0.002 | 0.78 (0.65,0.93) | 0.005 |
| 2-3 times per week | 0.75 (0.66,0.87) | 0.0001 | 0.76 (0.65,0.89) | 0.001 | 0.59 (0.50,0.70) | <1E-10 | 0.68 (0.57,0.82) | 4.5E-05 |
| 4+ times per week | 0.91 (0.77,1.08) | 0.285 | 1.12 (0.93,1.35) | 0.235 | 0.76 (0.62,0.94) | 0.010 | 1.01 (0.81,1.26) | 0.915 |
| **Smoking status** |  |  |  |  |  |  |  |  |
| Never-smoker | ref |  | ref |  | ref |  | ref |  |
| Ex-smoker | 1.16 (1.05,1.29) | 0.004 | 1.24 (1.11,1.38) | 0.0001 | 1.22 (1.07,1.38) | 0.002 | 1.29 (1.13,1.47) | 0.0001 |
| Current smoker | 1.94 (1.62,2.32) | <1E-10 | 1.76 (1.46,2.12) | 2.3E-09 | 1.79 (1.45,2.20) | <1E-10 | 1.66 (1.34,2.06) | 4.3E-06 |
| **Physical Activity Index (PAI)** |  |  |  |  |  |  |  |  |
| Inactive | ref |  | ref |  | ref |  | ref |  |
| Moderately inactive | 0.96 (0.85,1.09) | 0.545 | 0.86 (0.75,0.98) | 0.024 | 1.07 (0.91,1.25) | 0.423 | 0.97 (0.82,1.14) | 0.683 |
| Moderately active | 0.79 (0.70,0.89) | 0.0002 | 0.71 (0.62,0.81) | 1.9E-07 | 0.86 (0.74,1.01) | 0.059 | 0.83 (0.71,0.97) | 0.018 |
| Active | 0.71 (0.63,0.81) | <1E-10 | 0.6 (0.53,0.69) | <1E-10 | 0.85 (0.73,0.99) | 0.040 | 0.8 (0.68,0.94) | 0.006 |
| **Discrimination at work** |  |  |  |  |  |  |  |  |
| Not discriminated against at work | ref |  | ref |  | ref |  | ref |  |
| Discriminated against at work | 2.19 (1.99,2.40) | <1E-10 | 2.12 (1.92,2.33) | <1E-10 | 2.78 (2.50,3.10) | <1E-10 | 2.64 (2.35,2.95) | <1E-10 |
| **Redeployment** |  |  |  |  |  |  |  |  |
| Not redeployed | ref |  | ref |  | ref |  | ref |  |
| Redeployed | 1.24 (1.11,1.39) | 0.0002 | 1.15 (1.03,1.29) | 0.015 | 1.22 (1.07,1.40) | 0.003 | 1.17 (1.02,1.34) | 0.028 |
| Not working | 1.1 (0.96,1.26) | 0.155 | 0.95 (0.83,1.10) | 0.518 | 1.2 (1.02,1.40) | 0.024 | 1.09 (0.92,1.28) | 0.331 |
| **Job areas** |  |  |  |  |  |  |  |  |
| Not working in area specified | ref |  | ref |  | ref |  | ref |  |
| Ambulance | 1.18 (0.95,1.47) | 0.125 | 1.13 (0.90,1.42) | 0.291 | 1.02 (0.78,1.34) | 0.860 | 1.18 (0.89,1.57) | 0.250 |
| Community clinical | 0.86 (0.78,0.95) | 0.003 | 0.88 (0.80,0.98) | 0.019 | 0.82 (0.73,0.93) | 0.002 | 0.84 (0.74,0.95) | 0.006 |
| Community non-clinical | 1.22 (1.02,1.46) | 0.027 | 1.19 (0.99,1.43) | 0.061 | 1.16 (0.94,1.44) | 0.167 | 1.15 (0.93,1.43) | 0.200 |
| Emergency department | 1.22 (1.05,1.43) | 0.010 | 1.15 (0.97,1.35) | 0.104 | 1.12 (0.93,1.35) | 0.222 | 1.13 (0.93,1.38) | 0.209 |
| Intensive care | 1.39 (1.18,1.64) | 0.0001 | 1.19 (1.00,1.41) | 0.049 | 1.38 (1.13,1.67) | 0.001 | 1.28 (1.04,1.56) | 0.017 |
| Inpatient setting | 1.12 (1.02,1.24) | 0.022 | 1.05 (0.95,1.17) | 0.347 | 1.21 (1.07,1.36) | 0.002 | 1.21 (1.07,1.38) | 0.003 |
|  | Univariable |  | Adjusted<br>(Age, Sex, Ethnicity, Job role) |  | Univariable |  | Adjusted<br>(Age, Sex, Ethnicity, Job role) |  |
| Outpatient | 0.86 (0.77,0.97) | 0.010 | 0.94 (0.83,1.05) | 0.268 | 0.93 (0.81,1.07) | 0.303 | 1.04 (0.90,1.19) | 0.626 |
| Public / communal setting | 1.39 (1.08,1.80) | 0.012 | 1.36 (1.04,1.78) | 0.024 | 1.66 (1.24,2.21) | 0.001 | 1.76 (1.31,2.37) | 0.0001 |
| Mobile across areas | 1.03 (0.80,1.32) | 0.812 | 1 (0.78,1.30) | 0.974 | 1.24 (0.94,1.65) | 0.133 | 1.3 (0.97,1.73) | 0.077 |
| Prison | 1.07 (0.59,1.91) | 0.832 | 1.03 (0.57,1.87) | 0.931 | 1.19 (0.60,2.37) | 0.616 | 1.22 (0.61,2.45) | 0.570 |
| Other clinical setting | 1.03 (0.91,1.16) | 0.660 | 1.09 (0.96,1.24) | 0.164 | 1.3 (1.13,1.50) | 0.0002 | 1.37 (1.19,1.59) | 1.9E-05 |
| Nursing or care home | 0.92 (0.69,1.24) | 0.592 | 0.9 (0.67,1.22) | 0.500 | 1.21 (0.88,1.67) | 0.243 | 1.14 (0.82,1.58) | 0.445 |
| Psychiatric hospital / inpatient unit | 1.32 (1.02,1.71) | 0.035 | 1.26 (0.96,1.64) | 0.090 | 1.17 (0.85,1.61) | 0.325 | 1.13 (0.82,1.56) | 0.471 |
| At home | 1.06 (0.95,1.20) | 0.300 | 1.14 (1.01,1.28) | 0.041 | 0.89 (0.77,1.03) | 0.122 | 0.95 (0.81,1.10) | 0.483 |
| University | 1.06 (0.80,1.41) | 0.676 | 1.13 (0.84,1.50) | 0.418 | 0.85 (0.59,1.22) | 0.376 | 0.91 (0.62,1.32) | 0.605 |
| **Secure about raising concerns related to clinical practice** |  |  |  |  |  |  |  |  |
| Secure about raising concerns | ref |  | ref |  | ref |  | ref |  |
| Not secure in raising concerns | 2.08 (1.89,2.33) | <1E-10 | 2.04 (1.81,2.27) | <1E-10 | 2.22 (1.96,2.50) | <1E-10 | 2.08 (1.81,2.38) | <1E-10 |
| **Trusts in organisation to address concerns related to clinical practice** |  |  |  |  |  |  |  |  |
| Trusts organisation | ref |  | ref |  | ref |  | ref |  |
| Does not trust organisation | 1.89 (1.72,2.08) | <1E-10 | 1.89 (1.72,2.08) | <1E-10 | 1.92 (1.72,2.17) | <1E-10 | 1.89 (1.67,2.08) | <1E-10 |
| **Non-COVID-19 patient contact per week** |  |  |  |  |  |  |  |  |
| 0 | ref |  | ref |  | ref |  | ref |  |
| 1-5 | 1.07 (0.86,1.33) | 0.567 | 1 (0.80,1.25) | 0.100 | 0.76 (0.57,1.00) | 0.053 | 0.71 (0.54,0.95) | 0.021 |
| 6-20 | 0.94 (0.81,1.09) | 0.411 | 0.88 (0.75,1.03) | 0.099 | 0.77 (0.64,0.92) | 0.005 | 0.74 (0.61,0.89) | 0.002 |
| 21-50 | 0.96 (0.83,1.11) | 0.608 | 0.89 (0.76,1.04) | 0.132 | 0.94 (0.79,1.11) | 0.468 | 0.91 (0.76,1.09) | 0.297 |
| 51+ | 0.98 (0.85,1.13) | 0.753 | 0.95 (0.81,1.11) | 0.505 | 0.99 (0.84,1.18) | 0.931 | 0.99 (0.83,1.19) | 0.956 |
| **COVID-19 patient contact per week** |  |  |  |  |  |  |  |  |
| 0 | ref |  | ref |  | ref |  | ref |  |
| 1-5 | 1.14 (0.96,1.35) | 0.137 | 1.09 (0.92,1.30) | 0.332 | 1.37 (1.13,1.67) | 0.002 | 1.36 (1.11,1.67) | 0.003 |
| 6-20 | 1.17 (1.04,1.31) | 0.007 | 1.13 (1.00,1.28) | 0.048 | 1.31 (1.14,1.50) | 0.0001 | 1.33 (1.16,1.54) | 9.0E-05 |
| 21-50 | 1.46 (1.25,1.70) | <1E-10 | 1.3 (1.11,1.53) | 0.001 | 1.56 (1.30,1.88) | <1E-10 | 1.52 (1.26,1.85) | 1.6E-05 |
| 51+ | 2.11 (1.69,2.63) | <1E-10 | 1.74 (1.38,2.19) | 2.5E-06 | 2.33 (1.82,2.99) | <1E-10 | 2.08 (1.61,2.70) | 2.6E-08 |
| **Current working hours per week** | 1.01 (1.01,1.02) | <1E-10 | 1.01 (1.01,1.02) | 2.1E-05 | 1.01 (1.00,1.01) | 0.0003 | 1.01 (1.00,1.02) | 0.0003 |
| **Currently working at night** |  |  |  |  |  |  |  |  |
| No | ref |  | ref |  | ref |  | ref |  |
| Yes | 1.24 (1.13,1.37) | 0.00001 | 1.15 (1.03,1.28) | 0.012 | 1.27 (1.13,1.43) | 0.0001 | 1.25 (1.09,1.42) | 0.001 |
| **Access to appropriate PPE** |  |  |  |  |  |  |  |  |
| Not applicable or all the time | ref |  | ref |  | ref |  | ref |  |
| Some or most of the time | 1.66 (1.48,1.86) | <1E-10 | 1.56 (1.39,1.76) | <1E-10 | 1.84 (1.61,2.10) | <1E-10 | 1.78 (1.55,2.04) | <1E-10 |
| Rarely or not at all | 1.07 (0.65,1.76) | 0.799 | 1.27 (0.76,2.12) | 0.360 | 0.77 (0.38,1.53) | 0.455 | 0.82 (0.41,1.66) | 0.586 |

In multivariable analyses, participants belonging to the ‘Other’ ethnic group had higher odds of anxiety/depression compared to the White ethnic group (adjusted odds ratio (aOR) 1.33 [95%CI 0.98,1.80]). Odds ratios for the Asian (aOR 0.94 [95%CI 0.83,1.07]), Black (aOR 0.82 [95%CI 0.65,1.04]) and Mixed/multiple groups (aOR 0.95 [95%CI 0.76,1.18]) were slightly below 1. However, confidence intervals for these estimates were imprecise.

Asian (aOR 1.55 [95%CI 1.34,1.78]), Black (aOR 1.32 [95%CI 1.01,1.71]), Mixed/multiple (aOR 1.11 [95%CI 0.85,1.46]) and Other ethnic groups (aOR 1.83 [95%CI 1.29,2.60]) all had increased odds of PTSD compared to the White ethnic group. This association was particularly strong in Asian HCWs and HCWs belonging to the ‘Other’ ethnic group, however the latter was more imprecisely estimated.

Anxiety/depression and PTSD symptoms tended to share common predictors in our analysis. Higher odds were observed for both females and younger individuals, and all other job roles compared to doctors (with highest odds observed for nurses, nursing associates and midwives). Individuals living in areas of greater deprivation reported worse mental health, as did those who had been bereaved due to COVID-19 (own COVID-19 illness was not associated). Physical health factors were associated with mental health: participants who had ever smoked, and participants with long-term conditions reported worse mental health, and there was an apparent protective association with moderate or high levels of physical activity, and with moderate alcohol consumption. Higher levels of alcohol consumption were not protective.

Experiencing discrimination at work was one of the factors that showed the strongest association with poorer mental health (anxiety/depression: aOR 2.12 [95%CI 1.92,2.33]; PTSD: aOR 2.64 [95%CI 2.35,2.95]). Other strong associations were not feeling secure in raising concerns at work (anxiety/depression: aOR 2.04 [95%CI 1.81,2.27]; PTSD: aOR 2.08 [95%CI 1.81,2.38]), and not trusting one’s employer to address concerns (anxiety/depression: aOR 1.89 [95%CI 1.72,2.08]; PTSD: aOR 1.89 [95%CI 1.67,2.08]). Other work-related factors associated with both anxiety/depression and PTSD included working longer hours and reporting inadequate access to appropriate PPE. There was a dose-response association between contact with greater numbers of COVID-19 patients per week with both mental health measures, but seeing more patients without COVID-19 was not associated with increased odds.

Some predictor variables were more specific to one mental health outcome than the other: attributing at least some importance to religion was associated with higher odds of PTSD, but not with anxiety/depression. Living with a greater number of people was strongly associated with lower odds of anxiety/depression, but less strongly associated with PTSD.

Analyses were qualitatively similar when using non-imputed data (complete for confounders, **Supplementary Table 1**) and when including date of consent in the model (**Supplementary Table 2**). Participants consenting in January-March 2021 (compared to December 2020) generally had higher odds of poor mental health. Conclusions were qualitatively similar when additionally including participants who were not working at the time of baseline questionnaire completion (**Supplementary Table 3**).

### Associations of sociodemographic, physical health and work variables with ethnicity

**Table 3** presents associations between ethnicity and sociodemographic, physical health, and work factors, adjusted for age, sex and job role. These findings demonstrate the marked inequities by ethnicity across many factors associated with worse mental health.

**Table 3.** Association between ethnicity (as exposure, with White ethnic group as reference group) and multiple social, health and working factors (as separate outcomes), adjusted for age, sex and job role.

| Variable<br>(Coding)<sup>1</sup> | Model<sup>2</sup> | Asian/Asian British ethnic group |  | Black/Black British ethnic group |  | Mixed/Multiple ethnic group |  | Other ethnic group |  |
| --- | --- | --- | --- | --- | --- | --- | --- | --- | --- |
|  |  | Estimate<sup>2</sup> (95%CI) | P | Estimate<sup>2</sup> (95%CI) | P | Estimate<sup>2</sup> (95%CI) | P | Estimate<sup>2</sup> (95%CI) | P |
| Born in UK | Logistic | 0.102 (0.091, 0.115) | <1E-10 | 0.069 (0.056, 0.084) | <1E-10 | 0.483 (0.388, 0.601) | <1E-10 | 0.045 (0.032, 0.062) | <1E-10 |
| IMD decile<br>(Lower = more deprived) | Linear | -0.368 (-0.507, -0.229) | 2.29E-07 | -1.298 (-1.551, -1.044) | <1E-10 | -0.288 (-0.538, -0.038) | 0.024 | -0.816 (-1.171, -0.462) | 7.17E-06 |
| Religiosity<br>(‘Not at all’, to ‘Extremely’) | Ordered logistic | 4.948 (4.477, 5.469) | <1E-10 | 14.034 (11.766, 16.738) | <1E-10 | 1.219 (1.001, 1.484) | 0.049 | 7.18 (5.63, 9.159) | <1E-10 |
| Living with children | Logistic | 1.269 (1.144, 1.409) | 7.47E-06 | 1.282 (1.062, 1.547) | 0.010 | 1.034 (0.855, 1.25) | 0.731 | 1.299 (0.999, 1.688) | 0.051 |
| Living with adults 65+ | Logistic | 2.034 (1.691, 2.447) | <1E-10 | 1.268 (0.883, 1.822) | 0.199 | 1.393 (0.963, 2.016) | 0.078 | 1.666 (1.036, 2.681) | 0.035 |
| Bereaved due to COVID-19 | Logistic | 2.613 (2.351, 2.905) | <1E-10 | 2.52 (2.071, 3.066) | <1E-10 | 1.202 (0.997, 1.449) | 0.054 | 2.836 (2.159, 3.727) | <1E-10 |
| Long-term conditions<br>0, to 1, to 2 or more | Ordered logistic | 1.172 (1.032, 1.332) | 0.014 | 1.014 (0.8, 1.286) | 0.907 | 1.498 (1.211, 1.853) | 1.98E-04 | 1.028 (0.739, 1.43) | 0.869 |
| SARS-CoV-2 infection status | Logistic | 1.021 (0.904, 1.154) | 0.738 | 1.307 (1.059, 1.613) | 0.013 | 0.927 (0.74, 1.161) | 0.507 | 1.057 (0.778, 1.437) | 0.721 |
| Alcohol frequency<br>‘Never’ to ‘4+ times / week’ | Ordered logistic | 0.186 (0.168, 0.205) | <1E-10 | 0.178 (0.15, 0.211) | <1E-10 | 0.763 (0.647, 0.899) | 0.001 | 0.129 (0.1, 0.167) | <1E-10 |
| Smoking status<br>‘Never’, to ‘Ex’, to ‘Current’ | Ordered logistic | 0.481 (0.42, 0.552) | <1E-10 | 0.327 (0.245, 0.435) | <1E-10 | 1.265 (1.032, 1.551) | 0.023 | 0.862 (0.631, 1.178) | 0.352 |
| Physical Activity Index<br>‘Inactive’ to ‘Active’ | Ordered logistic | 0.829 (0.757, 0.909) | 6.04E-05 | 0.771 (0.655, 0.908) | 0.002 | 0.942 (0.796, 1.115) | 0.486 | 0.813 (0.641, 1.03) | 0.087 |
| Discriminated against at work | Logistic | 2.19 (1.958, 2.45) | <1E-10 | 3.162 (2.598, 3.849) | <1E-10 | 1.718 (1.404, 2.104) | 1.73E-07 | 2.303 (1.739, 3.05) | 7.52E-09 |
| Redeployed<sup>3</sup> | Logistic | 0.884 (0.772, 1.012) | 0.074 | 0.874 (0.683, 1.118) | 0.283 | 1.172 (0.934, 1.471) | 0.170 | 1.038 (0.749, 1.439) | 0.823 |
| Ambulance | Logistic | 0.186 (0.117, 0.295) | <1E-10 | 0.199 (0.081, 0.489) | 4.27E-04 | 0.464 (0.252, 0.852) | 0.013 | 0.467 (0.166, 1.318) | 0.150 |
| Community clinical | Logistic | 0.966 (0.860, 1.085) | 0.559 | 0.891 (0.717, 1.107) | 0.298 | 0.924 (0.748, 1.140) | 0.459 | 0.541 (0.385, 0.761) | 4.21E-04 |
| Community non-clinical | Logistic | 0.76 (0.587, 0.983) | 0.037 | 0.805 (0.505, 1.285) | 0.363 | 0.913 (0.605, 1.376) | 0.662 | 0.688 (0.319, 1.482) | 0.339 |
| Emergency department | Logistic | 1.138 (0.953, 1.358) | 0.152 | 1.202 (0.881, 1.639) | 0.245 | 1.068 (0.776, 1.469) | 0.688 | 1.228 (0.828, 1.820) | 0.307 |
| Intensive care | Logistic | 0.969 (0.791, 1.188) | 0.765 | 0.869 (0.589, 1.282) | 0.479 | 1.113 (0.787, 1.574) | 0.546 | 1.004 (0.612, 1.645) | 0.988 |
| Inpatient setting | Logistic | 1.139 (1.010, 1.285) | 0.034 | 1.011 (0.811, 1.260) | 0.922 | 0.921 (0.735, 1.154) | 0.473 | 1.734 (1.296, 2.321) | 2.13E-04 |
| Outpatient | Logistic | 1.168 (1.033, 1.321) | 0.013 | 0.83 (0.650, 1.061) | 0.136 | 0.921 (0.729, 1.164) | 0.492 | 1.352 (1.006, 1.817) | 0.045 |
| Public / communal setting | Logistic | 0.806 (0.588, 1.103) | 0.178 | 0.643 (0.334, 1.239) | 0.187 | 0.384 (0.168, 0.876) | 0.023 | 0.749 (0.341, 1.642) | 0.470 |
| Mobile across areas | Logistic | 0.596 (0.422, 0.841) | 0.003 | 0.593 (0.293, 1.201) | 0.146 | 0.768 (0.434, 1.359) | 0.365 | 0.964 (0.444, 2.090) | 0.926 |
|  |  | Estimate<sup>2</sup> (95%CI) | P | Estimate<sup>2</sup> (95%CI) | P | Estimate<sup>2</sup> (95%CI) | P | Estimate<sup>2</sup> (95%CI) | P |
| Other clinical setting | Logistic | 1.377 (1.201,1.578) | 4.57E-06 | 1.1 (0.850,1.423) | 0.470 | 0.834 (0.625,1.111) | 0.215 | 2.117 (1.571,2.852) | 8.22E-07 |
| Nursing or care home | Logistic | 0.691 (0.447,1.068) | 0.096 | 1.581 (0.935,2.675) | 0.088 | 0.715 (0.333,1.532) | 0.388 | 0.492 (0.120,2.010) | 0.323 |
| Psychiatric hospital / inpatient unit | Logistic | 1.228 (0.899,1.678) | 0.197 | 1.755 (1.089,2.827) | 0.021 | 1.505 (0.911,2.487) | 0.110 | 0.99 (0.429,2.285) | 0.981 |
| At home | Logistic | 0.916 (0.793,1.057) | 0.230 | 0.866 (0.657,1.142) | 0.307 | 1.155 (0.908,1.469) | 0.240 | 0.677 (0.449,1.019) | 0.061 |
| University | Logistic | 0.649 (0.459,0.920) | 0.015 | 0.589 (0.286,1.215) | 0.152 | 1.417 (0.867,2.316) | 0.164 | 0.817 (0.374,1.787) | 0.613 |
| Not secure about raising concerns related to clinical practice | Logistic | 1.678 (1.466,1.919) | <1E-10 | 1.698 (1.344,2.146) | 9.34E-06 | 0.943 (0.714,1.244) | 0.676 | 2.000 (1.460,2.740) | 1.64E-05 |
| Does not trust organisation to address concerns related to clinical practice | Logistic | 1.427 (1.279, 1.592) | 2.61E-10 | 1.074 (0.873, 1.319) | 0.500 | 1.161 (0.949, 1.425) | 0.147 | 1.357 (1.030, 1.789) | 0.030 |
| COVID-19 patient contact<br>(0, 1-5, 6-20, 21-50, 51+ pts/week) | Ordered logistic | 1.153 (1.04, 1.278) | 0.007 | 1.189 (0.991, 1.427) | 0.062 | 0.959 (0.794, 1.16) | 0.668 | 1.463 (1.139, 1.879) | 0.003 |
| Non-COVID-19 patient contact<br>(0, 1-5, 6-20, 21-50, 51+ pts/week) | Ordered logistic | 0.994 (0.906, 1.09) | 0.894 | 0.819 (0.691, 0.972) | 0.022 | 0.867 (0.734, 1.023) | 0.091 | 1.01 (0.796, 1.282) | 0.934 |
| Working hours | Linear | 0.246 (-0.243, 0.736) | 0.324 | 1.779 (0.898, 2.661) | 7.68E-05 | 0.104 (-0.772, 0.98) | 0.816 | 0.691 (-0.557, 1.938) | 0.278 |
| Night shift working | Logistic | 1.072 (0.947, 1.214) | 0.271 | 1.352 (1.09, 1.678) | 0.006 | 0.941 (0.748, 1.184) | 0.604 | 1.541 (1.147, 2.071) | 0.004 |
| Does not always have access to appropriate PPE<sup>3</sup> | Logistic | 1.629 (1.425, 1.863) | <1E-10 | 1.544 (1.219, 1.955) | 0.000 | 1.197 (0.929, 1.541) | 0.164 | 1.801 (1.316, 2.463) | 2.34E-04 |

<sup>1</sup>See also Table 1 and Supplementary Text. Regressions for ‘household size’ and ‘working in prison’ are not presented due to lack of model convergence
<sup>2</sup>Effect estimates are odds ratios (OR) for logistic and ordered logistic models, and beta coefficients for linear models
<sup>3</sup>Recoded to ‘yes’ vs ‘no’

We found more ethnic minority HCWs were living in the most deprived areas than White HCWs. Ethnic minority HCWs had twice to three times the odds of experiencing discrimination at work compared to White HCWs. Black HCWs worked longer hours, and were more likely to work night shifts (for HCWs in the Black or Other ethnic groups), have more contact with patients with COVID-19 (for HCWs in the Asian, Black or Other ethnic groups) and report reduced access to appropriate PPE. Ethnic minority HCWs were also less likely to feel secure raising concerns at work, or to have trust in their organisation to address these concerns. Participants from all ethnic minority groups were more likely than the White ethnic group to have been bereaved due to COVID-19, and Black HCWs were more likely to have had COVID-19 themselves. Protective associations were observed for some predictor variables in relation to mental health: overall, the odds of alcohol consumption were markedly lower in all HCWs from ethnic minority groups, and odds of smoking were lower in Asian and Black participants.

### Serial adjustment of associations between ethnicity and mental health

Serially adjusting for sociodemographic, physical health and work factors (**Figure 1, Supplementary Table 4**), attenuated the higher odds of PTSD for ethnic minority HCWs. After adjustment, the associations with anxiety/depression moved in a protective direction, with Asian and Black HCWs having reduced odds for these measures compared to White HCWs. Point estimates changed most after adjusting for the sociodemographic and work factors studied. Since this change in the pattern of results could potentially be driven by structural inequities in factors related to mental health that differ by ethnicity, ethnicity estimates for models 2-4 should not be interpreted as total effects, but as the effects after adjusting out inequities in these factors. Sensitivity analyses restricting to complete cases (N=6,278) showed similar results (**Supplementary Table 5**).

**Figure 1.**
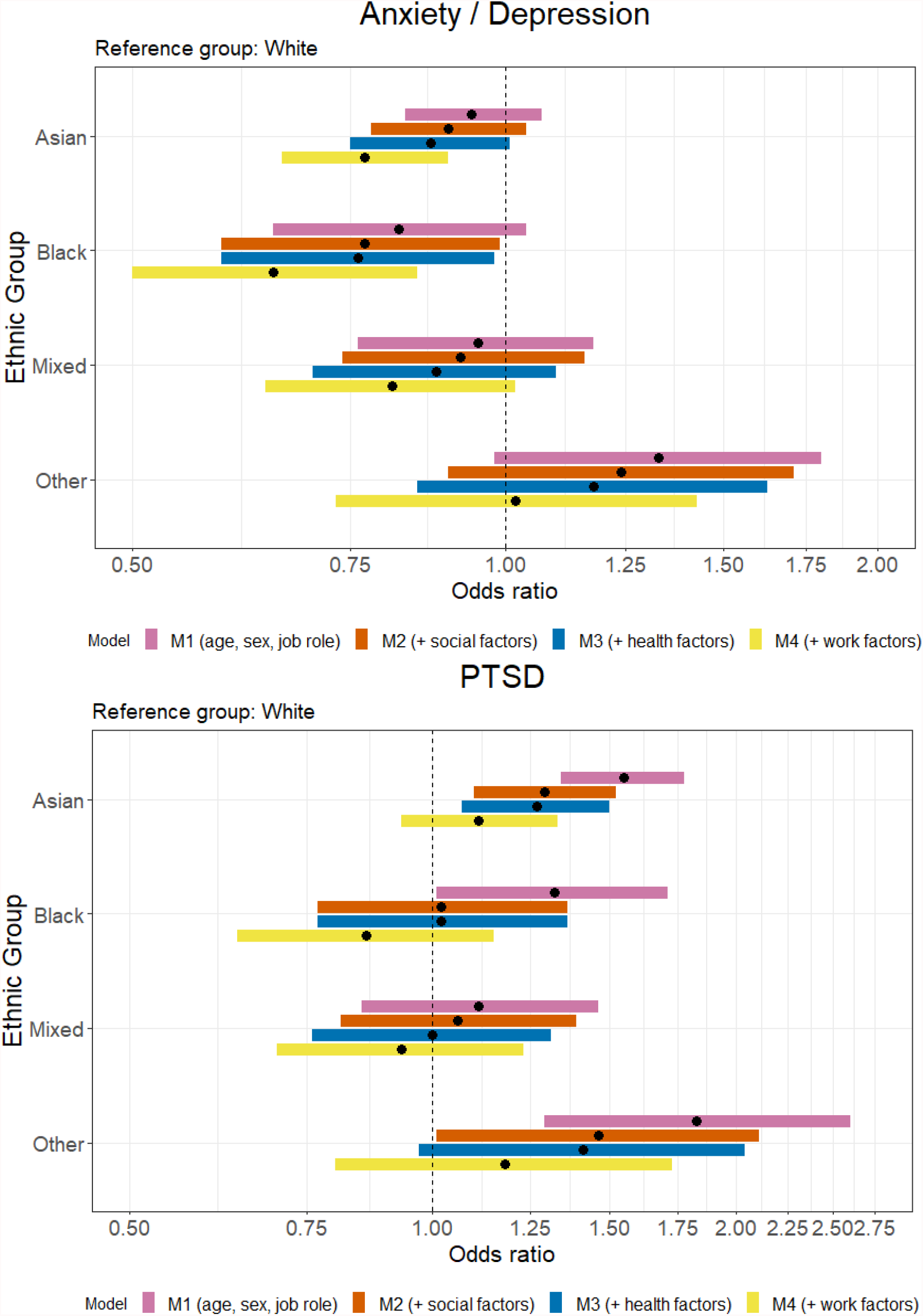
Odds ratios for each of four ethnic groups compared to the ‘White’ ethnic group for each of the four statistical models for Anxiety/Depression and PTSD.

## Discussion

In an analysis of almost 12,000 HCWs working in the UK, 1 in 4 met screening criteria for anxiety or depression, and 1 in 7 for PTSD. Odds of PTSD symptoms were higher in Asian, Black, Mixed/multiple and Other ethnic groups, compared to the White ethnic group. Differences in anxiety/depression symptoms by ethnicity were generally less pronounced, but suggested higher odds for the Other ethnic group and slightly lower odds for HCWs of Asian, Black and Mixed/multiple ethnicity compared to White HCWs.

Ethnic minority HCWs experienced disproportionately more factors associated with poorer mental health, including living in more deprived areas, workplace discrimination, working longer hours and working night shifts, seeing more COVID-19 patients, reporting reduced access to appropriate PPE, and being bereaved due to COVID-19. Younger, female participants, and those who were not doctors were also more likely to report anxiety/depression and PTSD symptoms. Serial adjustment for sociodemographic and work factors suggested that differences in mental health by ethnicity could be driven by structural inequities in these factors.

To our knowledge, this is one of the largest studies of mental health in HCWs to date. The overall sample size, and diversity of UK-REACH (∼30% from minority ethnic groups) enabled direct comparison of mental health symptoms by ethnicity.^22^ The baseline questionnaire collected data on many work, sociodemographic, physical and mental health factors, and follow-up questionnaires will enable the study of longitudinal changes in mental health.

Our study has limitations: in all cross-sectional studies, associations may be bidirectional, or due to reverse causation, e.g. smoking and mental health,^33^ religiosity and PTSD^34^. Moreover, although the breadth of UK-REACH data permitted adjustment for hypothesised confounders, residual confounding (due to missing data or measurement error) is possible in all observational studies.

Selection bias may also have affected our conclusions. Eligible individuals with worse mental health may be less likely to join a study^35^ and despite being similar to the NHS workforce in terms of age and sex,^36^ the current sample includes more clinical healthcare professionals,^36^ fewer ancillary workers, fewer participants from White and Black ethnic groups, and more from Asian and Mixed/multiple ethnic groups.^37^ Estimating prevalence of anxiety/depression and PTSD is difficult when using a screening tool and cut-offs to define caseness. Epidemiological studies may provide overoptimistic estimates if participants generally underreport symptoms, and biased associations may result from systematic variation in disclosure of symptom severity by levels or categories of a risk factor^38^. For example, under-reporting of symptoms by men has been hypothesised as a reason for why women may appear more susceptible to mental ill-health.^39^ Two studies of diverse participants reported that the PHQ-9 scale (from which the PHQ-2 is derived) performed similarly across ethnicities, suggesting that inequalities in symptoms were not due to differential performance of the scale by ethnicity.^40 41^

We used the five ONS ethnicity categories (rather than 18 ONS categories),^25^ due to small numbers in some groups. This limits our ability to capture differences in mental health for more granular categorisations of ethnicity. The ethnic categorisations used and quantitative associations presented cannot fully capture the nuance of participants’ lived experiences; however, complementary qualitative work in UK-REACH contextualises HCWs’ individual experiences, and also highlights the role of bereavement due to COVID-19, increased clinical workloads (including the traumatic and exhausting nature of treating high volumes of sick patients), and fear of contracting or transmitting infection (including PPE access) in contributing to mental ill-health in HCWs.^21^

Our finding that anxiety/depression symptoms were only weakly associated with ethnicity is similar to another study of HCW mental health during the pandemic;^5^ that study also found no strong associations between ethnicity and PTSD, which may reflect its smaller sample size and lower proportion of ethnic minority participants. However, our findings are similar to the results of the NHS Staff survey 2019,^42^ in which *lower* levels of pre-pandemic stress were generally reported by participants from Asian, Black, Chinese, and Mixed ethnic groups. Whilst stress is not directly comparable to our primary outcomes, before adjustment for age, sex, ethnicity and job role, we observed slightly lower odds of anxiety/depression for Asian and Black HCWs. This association weakened on adjustment, which may reflect that ethnic minority HCWs in UK-REACH are younger, and more likely to be doctors (younger age and being a doctor were associated with lower odds of mental ill-health in our analysis). Associations between mental health, age, sex and job role observed in our study align with other smaller quantitative studies in the UK,^5^ Kenya^43^ and the USA^44^ (the latter also confirmed associations with COVID-19-related workload, and overall working hours).

As in UK-REACH, smaller studies of HCWs in the UK have found strong associations between better mental health and feeling able to confide in one’s employer^45^ and between experiencing discrimination at work and probable anxiety/depression.^19^ The 2020 NHS Workforce Race Equality Standard (WRES) report^16^ found that experiencing discrimination was more common in HCWs from a minority ethnic background, and this association was similar in UK-REACH: all ethnic minority groups reported 2-3 times higher odds of workplace discrimination compared to White HCWs, with highest odds for Black HCWs. This highlights the need for preventative strategies and training to tackle discrimination in recruitment, retention, and performance appraisal, and the development of an inclusive culture, with clear policies for dealing with bullying and harassment (from staff, patients and public), supported by legislation^16^. As noted by the 2020 NHS WRES report, the pandemic has not created inequity, but has “thrown it into sharp relief”.^16^ We suggest the same is true for many other workplace stressors associated with mental ill-health, experienced disproportionately by ethnic minority HCWs. Furthermore, multiple areas of inequity or marginalisation may intersect.^46^ Some factors such as bereavement, both personal and collective, have been exacerbated by the pandemic.^47^ This underlines the importance of employers being aware of communities most affected, and of compassion and flexibility (e.g. via provision of culturally sensitive bereavement support^48^) to all staff who have lived through traumatic experiences,^49^ inside and outside of work. The National Institute for Clinical Excellence (NICE) recognises being a HCW, exposure to a traumatic event, multiple life stressors, and social disadvantage as risk factors for PTSD.^50^ This would be consistent with our findings that HCWs’ experiencing intersecting stressors at work and in their home lives are more likely to have PTSD symptoms. Attenuation of associations between ethnicity and PTSD observed after adjusting for multiple stressors may help evaluate the magnitude of disparity in mental health by ethnicity that would remain if these intermediate or downstream factors were intervened upon. Multiple agencies have underlined the value of considering specialist trauma services designed to support other professions (e.g. the military) when designing services for HCWs.^51^ Any workplace support service must be accompanied by truly protected time for all staff (working different shift patterns) to utilise them.

Although cross-sectional studies cannot infer causality, understanding associations between workplace factors and poor mental health may inform future work to develop proactive and reactive strategies for safeguarding the mental health of ethnic minority HCWs and the healthcare workforce as a whole. Multiple factors associated with worse mental health in HCWs were experienced disproportionately by ethnic minority HCWs. Future work will focus on determining the longitudinal impact of risk factors on mental health using follow-up questionnaires in UK-REACH, and on analysing other hypothesised risk factors not recorded at baseline, such as moral injury and burnout.^6^

There is a moral imperative of caring for HCWs’ wellbeing both during the COVID-19 pandemic and beyond, which includes tackling structural inequities at work; predictions of workforce attrition also underline the practical imperative of doing so.^52^ Prevailing discourses of being ‘at war’ with COVID-19, and the narrative of heroism, have been criticised as dehumanising, and silencing of HCWs’ concerns about working conditions and mental health, thus disenfranchising their collective anger, grief^53^ and exhaustion.^52^ Listening to, and learning from the lived experiences of the diverse healthcare workforce contending with extraordinarily challenging working conditions is crucial to inform the development of compassionate, culturally sensitive^48^ workplace policies to support HCW wellbeing and mental health. However, considering the pressures on the entire workforce, policymakers must acknowledge that sustainable improvements in staff wellbeing will only be possible alongside investment in NHS recruitment, retention and fair remuneration, to ensure adequate staffing.^54^ Together with concerted efforts by employers to address workplace inequity/discrimination, such strategies may help improve working conditions and HCW mental health and wellbeing.

## Supporting information

STROBE_checklist

## Data Availability

Deidentified participant data are available via a system of managed access, to access data or samples produced by the UK-REACH study please contact to discuss your request. The protocol for the UK-REACH longitudinal cohort study is available at: http://dx.doi.org/10.1136/bmjopen-2021-050647, and a Data Dictionary of the baseline questionnaire data is available at: https://uk-reach.org/main/data-dictionary/.

## Funding

UK-REACH is supported by a grant (MR/V027549/1) from the MRC-UK Research and Innovation (UKRI) and the Department of Health and Social Care through the National Institute for Health Research (NIHR) rapid response panel to tackle COVID-19. Core funding was also provided by NIHR Biomedical Research Centres. ALG was funded by internal fellowships at the University of Leicester from the Wellcome Trust Institutional Strategic Support Fund (204801/Z/16/Z) and the BHF Accelerator Award (AA/18/3/ 34220). LBN is supported by an Academy of Medical Sciences Springboard Award (SBF005\1047). CJ was funded by a Medical Research Council Clinical Research Training Fellowship (MR/P00167X/1). CAM is an NIHR Academic Clinical Fellow (ACF-2018-11-004). MDT holds a Wellcome Trust Investigator Award (WT 202849/Z/ 16/Z) and an NIHR Senior Investigator Award. KW is funded through an NIHR Career Development Fellowship (CDF-2017-10-008). MP is funded by an NIHR Development and Skills Enhancement Award and with KK, also acknowledges support from the NIHR Leicester BRC and with KK and LJG, acknowledges support from NIHR ARC East Midlands. LBN is supported by the Academy of Medical Sciences (SBF005/1047). This work is carried out with the support of BREATHE—the Health Data Research Hub for Respiratory Health (MC_PC_19004) funded through the UK Research and Innovation Industrial Strategy Challenge Fund and delivered through Health Data Research UK. For the purpose of open access, the author has applied a CC BY public copyright licence to any Author Accepted Manuscript version arising from this submission.

## Acknowledgements

We would like to thank all the healthcare workers who have taken part in this study whilst the NHS is under immense pressure.

Some of the questionnaire items in the UK-REACH questionnaire are taken or adapted from questionnaire material developed as part of the Longitudinal Population Studies COVID-19 questionnaire. This was supported by Wellcome “Longitudinal Population Study Covid-19 Steering Group and Secretariat” as a Strategic Support Science Grant - 221574/Z/20/Z.

We wish to acknowledge the Professional Expert Panel group (Amir Burney, Association of Pakistani Physicians of Northern Europe; Tiffanie Harrison; London North West University Healthcare NHS Trust; Ahmed Hashim, Sudanese Doctors Association; Sandra Kazembe, University Hospitals Leicester NHS Trust; Susie M. Lagrata (Co-chair), Filipino Nurses Association, UK & University College London Hospitals NHS Foundation Trust; Satheesh Mathew, British Association of Physicians of Indian Origin; Juliette Mutuyimana, Kingston Hospitals NHS Trust; Padmasayee Papineni (Co-chair), London North West University Healthcare NHS Trust; Tatiana Monteiro, University Hospitals Leicester NHS Trust),the UK-REACH Stakeholder Group (see the cohort study protocol^55^ for details), the Study Steering Committee, SERCO, and the following people for their support in setting up the study from the regulatory bodies: Kerrin Clapton and Andrew Ledgard (General Medical Council), Caroline Kenny (Nursing and Midwifery Council), David Teeman and Lisa Bainbridge (General Dental Council), My Phan and Jenny Clapham (General Pharmaceutical Council), Angharad Jones (General Optical Council), Katherine Timms and Charlotte Rogers (The Health and Care Professions Council) and Mark Neale (Pharmaceutical Society of Northern Ireland).

We would also like to acknowledge the following trusts and sites who recruited participants to the study: Nottinghamshire Healthcare NHS Foundation Trust, University Hospitals Leicester, Lancashire Teaching Hospitals NHS Foundation Trust, Northumbria Healthcare, Berkshire Healthcare, Derbyshire Healthcare NHS Foundation Trust, South Tees NHS Foundation Trust, Birmingham and Solihull NHS Foundation Trust, Affinity Care, Royal Brompton and Harefield, Sheffield Teaching Hospitals, St George’s Hospital, Yeovil District Hospital, Lewisham and Greenwich NHS Trust, Black Country Community Healthcare NHS Foundation Trust, Sussex Community NHS Foundation Trust, South Central Ambulance Service, University Hospitals Coventry and Warwickshire, University Hospitals Southampton NHS Foundation Trust, London Ambulance Trust, Royal Free, Birmingham Community Healthcare NHS Foundation Trust, Central London Community Healthcare, Chesterfield Royal Hospital, Bridgewater Community Healthcare, Northern Borders, County Durham and Darlington Foundation Trust, Walsall Healthcare NHS Trust.

## Notes

### Clinical Protocols

http://dx.doi.org/10.1136/bmjopen-2021-050647

## References

1. Office for National Statistics. Healthcare in the United Kingdom: UK Government; 2021 [Available from: https://coronavirus.data.gov.uk/details/healthcare accessed 27th October 2021 2021.

2. NHS Digital. Appointments in General Practice (Official statistics, Experimental statistics): NHS Digital; 2021 [Available from: <https://digital.nhs.uk/data-and-information/publications/statistical/appointments-in-general-practice> accessed 27th October 2021 2021.

3. Mahase E. Covid-19: Hospitals in crisis as ambulances queue and staff are asked to cancel leave. BMJ 2020;371:m4980. doi: 10.1136/bmj.m4980

4. Davies G. The supply of personal protective equipment (PPE) during the COVID-19 pandemic. In: National Audit Office, ed. London, UK, 2020.

5. Lamb D, Gnanapragasam S, Greenberg N, et al. Psychosocial impact of the COVID-19 pandemic on 4378 UK healthcare workers and ancillary staff: initial baseline data from a cohort study collected during the first wave of the pandemic. Occup Environ Med 2021 doi: 10.1136/oemed-2020-107276 [published Online First: 2021/06/30]

6. French L, Hanna P, Huckle C. “If I die, they do not care”: U.K. National Health Service staff experiences of betrayal-based moral injury during COVID-19. Psychological Trauma: Theory, Research, Practice, and Policy 2021:No Pagination Specified-No Pagination Specified. doi: 10.1037/tra0001134

7. Wanigasooriya K, Palimar P, Naumann DN, et al. Mental health symptoms in a cohort of hospital healthcare workers following the first peak of the COVID-19 pandemic in the UK. BJPsych Open 2021;7(1):e24. doi: 10.1192/bjo.2020.150 [published Online First: 2020/12/29]

8. d’Ettorre G, Ceccarelli G, Santinelli L, et al. Post-Traumatic Stress Symptoms in Healthcare Workers Dealing with the COVID-19 Pandemic: A Systematic Review. Int J Environ Res Public Health 2021;18(2) doi: 10.3390/ijerph18020601 [published Online First: 2021/01/16]

9. Mansfield KE, Mathur R, Tazare J, et al. Indirect acute effects of the COVID-19 pandemic on physical and mental health in the UK: a population-based study. The Lancet Digital Health 2021;3(4):e217–e30. doi: https://doi.org/10.1016/S2589-7500(21)00017-0

10. Chattopadhyay I, Davies G, Adhiyaman V. The contributions of NHS healthcare workers who are shielding or working from home during COVID-19. Future Healthcare Journal 2020;7(3):e57–e59. doi: 10.7861/fhj.2020-0096

11. Office for National Statistics. Coronavirus (COVID-19) related deaths by ethnic group, England and Wales: 2 March 2020 to 15 May 2020: UK Government; 2020 [Available from: https://www.ons.gov.uk/peoplepopulationandcommunity/birthsdeathsandmarriages/deaths/articles/coronaviruscovid19relateddeathsbyethnicgroupenglandandwales/2march2020to15may2020 accessed 27th October 2021 2021.

12. Office for National Statistics. Deaths involving the coronavirus (COVID-19) among health and social care workers in England and Wales, deaths registered between 9 March and 20 July 2020: UK Government; 2021 [Available from: https://www.ons.gov.uk/peoplepopulationandcommunity/healthandsocialcare/causesofdeath/adhocs/12112deathsinvolvingthecoronaviruscovid19amonghealthandsocialcareworkersinenglandandwalesdeathsregisteredbetween9marchand20july2020 accessed 27th October 2021 2021.

13. Nafilyan V, Islam N, Mathur R, et al. Ethnic differences in COVID-19 mortality during the first two waves of the Coronavirus Pandemic: a nationwide cohort study of 29 million adults in England. European Journal of Epidemiology 2021;36(6):605–17. doi: 10.1007/s10654-021-00765-1

14. Cook T, Kursumovic E, Lennane S. Exclusive: deaths of NHS staff from covid-19 analysed. Health Service Journal 2020.

15. Martin C, Pan D, Melbourne C, et al. Predictors of SARS-CoV-2 infection in a multi-ethnic cohort of United Kingdom healthcare workers: a prospective nationwide cohort study (UK-REACH)2021.

16. WRES Implementation team. NHS Workforce Race Equality Standard: 2020 Data Analysis Report for NHS Trusts and Clinical Commissioning Groups, 2021.

17. Marmot M, Allen J, Goldblatt P, et al. Build Back Fairer: The COVID-19 Marmot Review. The Pandemic, Socioeconomic and Health Inequalities in England. London, UK: Institute of Health Equity, 2020.

18. NHS Staff Surveys. NHS Staff Survey National Response [Data]. 2021 NHS Staff Survey; 2020 [Available from: https://www.nhsstaffsurveys.com/static/52cd823a2002538cc2f65f125bc578e2/NHS-Staff-Survey-2020-National-WRES-WDES-data_v2.xlsx accessed 27th October 2021 2021.

19. Rhead RD, Chui Z, Bakolis I, et al. Impact of workplace discrimination and harassment among National Health Service staff working in London trusts: results from the TIDES study. BJPsych Open 2020;7(1):e10. doi: 10.1192/bjo.2020.137 [published Online First: 2020/12/17]

20. Uphoff EP, Lombardo C, Johnston G, et al. Mental health among healthcare workers and other vulnerable groups during the COVID-19 pandemic and other coronavirus outbreaks: A rapid systematic review. PLoS One 2021;16(8):e0254821. doi: 10.1371/journal.pone.0254821 [published Online First: 2021/08/05]

21. Qureshi I, Gogoi M, Al-Oraibi A, et al. Mental Health During COVID-19: A Qualitative Study with Ethnically Diverse Healthcare Workers in the United Kingdom. medRxiv 2021:2021.12.13.21267718. doi: 10.1101/2021.12.13.21267718

22. Woolf K, Melbourne C, Bryant L, et al. The United Kingdom Research study into Ethnicity And COVID-19 outcomes in Healthcare workers (UK-REACH): protocol for a prospective longitudinal cohort study of healthcare and ancillary workers in UK healthcare settings. BMJ Open 2021;11(9):e050647. doi: 10.1136/bmjopen-2021-050647

23. Kroenke K, Spitzer RL, Williams JB, et al. An ultra-brief screening scale for anxiety and depression: the PHQ-4. Psychosomatics 2009;50(6):613–21. doi: 10.1176/appi.psy.50.6.613 [published Online First: 2009/12/10]

24. Lang AJ, Stein MB. An abbreviated PTSD checklist for use as a screening instrument in primary care. Behav Res Ther 2005;43(5):585–94. doi: 10.1016/j.brat.2004.04.005 [published Online First: 2005/05/04]

25. Office for National Statistics. 2011 Census Variable and Classification Information: Part 4. 2011 Census: User Guide, 2014.

26. Ministry of Housing CaLG. English Indices of Deprivation 2019. 2019 [Available from: https://www.gov.uk/government/statistics/english-indices-of-deprivation-2019 accessed 2nd November 2021.

27. Physical Activity Policy HID. The General Practice Physical Activity Questionnaire (GPPAQ): A screening tool to assess adult physical activity levels, within primary care London, UK, 2009.

28. Rubin D. Multiple Imputation for Nonresponse in Surveys: John Wiley & Sons, Inc. 1987.

29. Sterne JA, Davey Smith G. Sifting the evidence-what’s wrong with significance tests? Bmj 2001;322(7280):226–31. doi: 10.1136/bmj.322.7280.226 [published Online First: 2001/02/13]

30. Wasserstein RL, Lazar NA. The ASA Statement on p-Values: Context, Process, and Purpose. The American Statistician 2016;70(2):129–33. doi: 10.1080/00031305.2016.1154108

31. Goudswaard LJ, Harrison S, Van De Klee D, et al. Blood pressure variability and night-time dipping assessed by 24-hour ambulatory monitoring: Cross-sectional association with cardiac structure in adolescents. PLoS One 2021;16(6):e0253196. doi: 10.1371/journal.pone.0253196 [published Online First: 2021/06/17]

32. StataCorp LLC. Stata Statistical Software: Release 16. College Station, TX, USA., 2019.

33. Treur JL, Munafò MR, Logtenberg E, et al. Using Mendelian randomization analysis to better understand the relationship between mental health and substance use: a systematic review. Psychol Med;51(10):1593–624. doi: 10.1017/s003329172100180x

34. Slater CL, Bordenave J, Boyer BA. Impact of Spiritual and Religious Coping on PTSD. In: Martin CR, Preedy VR, Patel VB, eds. Comprehensive Guide to Post-Traumatic Stress Disorders. Cham: Springer International Publishing 2016:147–62.

35. Howe LD, Tilling K, Galobardes B, et al. Loss to Follow-up in Cohort Studies: Bias in Estimates of Socioeconomic Inequalities. Epidemiology 2013;24(1):1–9. doi: 10.1097/EDE.0b013e31827623b1

36. NHS Digital. NHS Workforce Statistics - April 2021 (Including selected provisional statistics for May 2021). In: NHS Digital, ed. NHS Workforce Statistics, 2021.

37. NHS Employers. Ethnicity in the NHS Infographic 2019 [updated 12 May 2019. Available from: https://www.nhsemployers.org/articles/ethnicity-nhs-infographic accessed 13th September 2021.

38. Kessler RC. Psychiatric epidemiology: selected recent advances and future directions. Bull World Health Organ 2000;78(4):464–74. [published Online First: 2000/07/08]

39. Smith DT, Mouzon DM, Elliott M. Reviewing the Assumptions About Men’s Mental Health: An Exploration of the Gender Binary. Am J Mens Health 2018;12(1):78–89. doi: 10.1177/1557988316630953 [published Online First: 2016/02/13]

40. Patel JS, Oh Y, Rand KL, et al. Measurement invariance of the patient health questionnaire-9 (PHQ-9) depression screener in U.S. adults across sex, race/ethnicity, and education level: NHANES 2005-2016. Depress Anxiety 2019;36(9):813–23. doi: 10.1002/da.22940 [published Online First: 2019/07/30]

41. Galenkamp H, Stronks K, Snijder MB, et al. Measurement invariance testing of the PHQ-9 in a multi-ethnic population in Europe: the HELIUS study. BMC Psychiatry 2017;17(1):349. doi: 10.1186/s12888-017-1506-9 [published Online First: 2017/10/27]

42. NHS Staff Survey. NHS Staff Survey 2019: Survey Coordination Centre, 2019.

43. Shah J, Monroe-Wise A, Talib Z, et al. Mental health disorders among healthcare workers during the COVID-19 pandemic: a cross-sectional survey from three major hospitals in Kenya. BMJ Open 2021;11(6):e050316. doi: 10.1136/bmjopen-2021-050316 [published Online First: 2021/06/11]

44. Bryant-Genevier J, Rao CY, Lopes-Cardozo B, et al. Symptoms of Depression, Anxiety, Post-Traumatic Stress Disorder, and Suicidal Ideation Among State, Tribal, Local, and Territorial Public Health Workers During the COVID-19 Pandemic - United States, March-April 2021. MMWR Morb Mortal Wkly Rep 2021;70(26):947–52. doi: 10.15585/mmwr.mm7026e1 [published Online First: 2021/07/02]

45. Greene T, Harju-Seppänen J, Adeniji M, et al. Predictors and rates of PTSD, depression and anxiety in UK frontline health and social care workers during COVID-19. Eur J Psychotraumatol 2021;12(1):1882781. doi: 10.1080/20008198.2021.1882781 [published Online First: 2021/05/11]

46. Qureshi I, Gogoi M, Al-Oraibi A, et al. Intersectionality and developing evidence-based policy. Lancet 2022;399(10322):355–56. doi: 10.1016/s0140-6736(21)02801-4 [published Online First: 2022/01/24]

47. Kumar RM. The Many Faces of Grief: A Systematic Literature Review of Grief During the COVID-19 Pandemic. Illness, Crisis & Loss 2021:10541373211038084. doi: 10.1177/10541373211038084

48. National Health and Wellbeing Team NEaNI. Our NHS People Understanding different bereavement practices and how our colleagues may experience grief 2020 [17th November 2021]. Available from: https://www.eastlondonhcp.nhs.uk/downloads/End%20of%20Life%20Care/Bereavement%20Practices.pdf.

49. Mind. Our Frontline: support for healthcare workers., 2021.

50. Excellence NIfC. Post-traumatic stress disorder: What are the risk factors? 2020 [17th November 2021]. Available from: <https://cks.nice.org.uk/topics/post-traumatic-stress-disorder/background-information/risk-factors/>.

51. Rimmer A. Covid-19: Offer staff military-style mental health support, say healthcare leaders. BMJ 2021;373:n1292. doi: 10.1136/bmj.n1292

52. Sheather J, Slattery D. The great resignation—how do we support and retain staff already stretched to their limit? BMJ 2021;375:n2533. doi: 10.1136/bmj.n2533

53. Maddrell A. Bereavement, grief, and consolation: Emotional-affective geographies of loss during COVID-19. Dialogues in Human Geography 2020;10(2):107–11. doi: 10.1177/2043820620934947

54. Quirk H, Crank H, Carter A, et al. Barriers and facilitators to implementing workplace health and wellbeing services in the NHS from the perspective of senior leaders and wellbeing practitioners: a qualitative study. BMC Public Health 2018;18(1):1362. doi: 10.1186/s12889-018-6283-y

55. Woolf K, Melbourne C, Bryant L, et al. The United Kingdom Research study into Ethnicity And COVID-19 outcomes in Healthcare workers (UK-REACH): Protocol for a prospective longitudinal cohort study of healthcare and ancillary workers in UK healthcare settings. medRxiv 2021:2021.02.23.21251975. doi: 10.1101/2021.02.23.21251975

