## Supplementary_information for "Mental health in a diverse sample of healthcare workers during the COVID-19 pandemic: cross-sectional analysis of the UK-REACH study"

**Supplementary Text.** Details of how individual variables were measured and coded for analysis.

| **Variable** | **Description** |
| --- | --- |
| **Age** | Continuous variable. Age in years. Derived from date of birth entered by participants at registration. |
| **Sex** | Binary variable. Participants were asked their sex assigned at birth. |
| **Ethnicity** | Categorical variable. Participants were asked to select their ethnicity from a list of the 18 Office for National Statistics categories. These were then combined into five categories as per the 2011 Census, as follows:    **Asian**  Asian/Asian British – Indian  Asian/Asian British – Pakistani  Asian/Asian British – Bangladeshi  Asian/Asian British – Chinese  Asian/Asian British - Any other Asian background    **Black**  Black/African/Caribbean/Black British  - African  Black/African/Caribbean/Black British – Caribbean  Black/African/Caribbean/Black British - Any other Black/African/Caribbean background    **Mixed**  Mixed/Multiple ethnic groups - White and Black Caribbean  Mixed/Multiple ethnic groups - White and Black African  Mixed/Multiple ethnic groups - White and Asian  Mixed/Multiple ethnic groups - Any other Mixed/multiple ethnic background    **White**  White - English/Welsh/Scottish/Northern Irish/British  White – Irish  White - Gypsy or Irish Traveller  White - Any other white background    **Other**  Other ethnic group – Arab  Other ethnic group - Any other ethnic background |
| **Job** | Categorical variable. Participants were asked to select their main job/role. Categorised as below:    **Doctor or medical support** **–** Doctor, Advanced Critical Care Practitioner, Anaesthesia associate, Physician associate, Surgical Care Practitioner, Other medical associate  **Nurse, NA or Midwife –** Advanced Nurse Practitioner, Healthcare assistant, Maternity support worker, Midwife, Nurse, Nursing Associate (NA), Other nursing and midwifery role,  **Allied Health Professional (including pharmacists, ambulance workers and those in optical roles) –** Arts therapist, Biomedical scientist, Chiropodist/Podiatrist, Clinical scientist, Dietician, Hearing aid dispenser, Occupational therapist, Operating department practitioner, Orthoptist, Physiotherapist, Practitioner psychologist, Prosthetist/Orthotist, Radiographer, Speech and language therapist, Other Allied Health Professional role, Emergency medical, Paramedic, Other ambulance role, OT Support, Phlebotomist, Physiotherapy Assistant, Radiography Assistant, Other clinical support role, Pharmacist , Pharmacy technician, Other pharmacy role, Optical - Dispensing optician, Optometrist, Other Optical role  **Dental –** Clinical dental technician, Dental Hygienist, Dental nurse, Dental technician, Dentist, Other dental role  **Admin, estates or other –** Administration, Catering services, Domestic services, Estates services, Porter, Other wider healthcare role, Any other role |
| **Migration status** | Binary variable. Participants were asked whether they were born in the UK. |
| **Index of Multiple Deprivation (IMD) quintile** | Ordinal variable. Participants provided their residential postcode on registration for the study. This was used to determine the Index of Multiple Deprivation (the official measure of deprivation for small areas of England) in the area in which they live. The IMD ranks all areas in England based on 7 measures of deprivation and the ranks can be expressed as quintiles. Lower quintiles indicate more deprivation. Although Wales, Scotland and Northern Ireland have their own measures of deprivation, these are said not to be directly comparable to English IMD and therefore we elected to impute an ‘English IMD’ for residents of the these nations. |
| **Household size** | Continuous variable. Participants were asked how many people live in their house other than themselves. |
| **Religiosity** | Ordinal variable. Participants were asked “How important is religion to you in your everyday life?” and could answer using the following scale: Not at all important, Fairly important, Very important, Extremely important and Prefer not to answer. This question was asked only to those who indicated that they identified as belonging to a particular religious group in a previous question. Those who indicated they had ‘No religion’ were grouped together with those indicating religion was ‘Not at all important’. |
| **Living with children** | Binary variable. Participants were asked the ages of the individuals they live with. The age for each individual was categorised as: 0-1, 2-4, 5-10, 11-16, 17-18, 19-24, 25-34, 35-44, 45-54, 55-64, 65-74, 75-84, 85-94, 95+. Living with children was defined as participants living with individuals aged 18 or below. |
| **Living with adults 65+** | Binary variable. Participants were asked the ages of the individuals they live with as above. Any option chosen from 65-74 upwards was used to identify a participant that lived with adults aged 65+. |
| **Bereavement due to COVID-19** | Binary variable. Participants were asked “Do you personally know anyone who has died from COVID-19 (not including patients you have cared for as part of your work)?“. Participants who answered ‘Yes’ to any of the options (‘family members’,’friends’,’colleagues’, ’Yes, someone else’ ) were categorised as ‘Knows someone who died’. |
| **Long-term conditions** | Ordinal variable. Participants were asked to select whether any of the following applied to them: ‘Organ transplant’, ‘Diabetes (Type I or II)’, ‘Heart disease or heart problems’, ‘Stroke’, ‘Kidney disease’, ‘Liver disease’, ‘Asthma’, ‘Other lung condition such as COPD, bronchitis or emphysema’, ‘Cancer’, ‘Condition affecting the brain and nerves (e.g. Dementia, Parkinson’s, Multiple Sclerosis)’, ‘A weakened immune system or reduced ability to deal with infections (as a result of a disease or treatment)’. |
| **SARS-CoV-2 infection** | Binary variable. Participant was categorised as having had a ‘COVID-19 infection’ if they reported a positive swab or antibody test, or if they thought they had had COVID-19 when asked ‘Do you think that you currently have or have had COVID-19?’ |
| **Alcohol Frequency** | Categorical variable. Participants were asked ‘How often do you have a drink containing alcohol?’ |
| **Smoking status** | Categorical variable. Participants were asked to indicate their current smoking status. The categorisations were: ‘Current smoker’, ‘Ex-smoker’, ‘Never smoker’. |
| **Physical Activity Index (PAI)** | Categorical variable. 4-level variable derived from the General Practice Physical Activity Questionnaire (GPPAQ) included in the questionnaire. Individuals were categorised using responses to questions: ‘Think about a typical week at work over the past month. Please consider the type and amount of physical activity involved in your work. Please select one option only’, and, ‘During the last week, about how many hours did you spend on each of the following activities’ – responses for: ‘Physical exercise such as swimming, jogging, aerobics, football, tennis, gym workout etc.’, and, ‘Cycling, including cycling to work and during leisure time’ were used to categorise individuals (as ‘Walking’, ‘housework/childcare’ and ‘gardening/DIY’ not sufficiently reliable for understanding activity levels) – for reference see: https://psnc.org.uk/suffolk-lpc/wp-content/uploads/sites/108/2019/05/GPPAQ.pdf |
| **Discrimination at work** | Binary variable. Participants were asked: ‘In the last 12 months have you personally experienced discrimination at work from any of the following? Select all that apply.’ Participants were categorised as ‘Discriminated against at work’ if they selected the options ‘Patients / service users, their relatives or other members of the public’, or ‘Manager / team leader or other colleagues’. |
| **Redeployment** | Categorical variable. Participants were asked: ‘During the UK national lockdown that began on 23rd March 2020, were you redeployed to a different role because of the pandemic?’. Those who were not working at the time were categorised as ‘Not working’, thus providing three categories: ‘Not redeployed’, ‘Redeployed’ and ‘Not working’. |
| **Job areas** | Binary variables. Participants were asked: ‘Please indicate which areas you work in a typical week now’. This was a list of non-mutually exclusive clinical and non-clinical areas. |
| **Secure about raising concerns** **related to clinical practice** | Binary variable. Participants were asked how much they agreed with the statement: ‘I would feel secure raising concerns about unsafe clinical practice’. Participants were categorised as ‘Secure in raising concerns ’ if they responded with ‘Agree’ or ‘Strongly agree’. |
| **Trust in organisation to address concerns related to clinical practice** | Binary variable. Participants were asked how much they agreed with the statement: ’I am confident that my organisation would address my concern.’ Participants were categorised as ‘Trusts organisation’ if they responded with ‘Agree’ or ‘Strongly agree’. |
| **COVID-19 patient contact per week** | Categorical variable. Derived from combining the midpoints of responses (0,1-5,6-20,21-50,51+) to the number of ‘confirmed or suspected COVID-19’ patients talked with at work last week, both ‘face to face with social distancing’ and ‘with physical contact’. Midpoints were then added together and groups were then categorised as before (0,1-5,6-20,21-50,51+). For the ‘51+’ option where there was no midpoint, ‘51’ was used for the purposes of response summation. |
| **Non-COVID-19 patient contact per week** | Categorical variable. Derived from combining the midpoints of responses (0,1-5,6-20,21-50,51+) to the number of patients talked with ‘remotely’ or ‘other patients’ talked with at work last week both ‘face to face with social distancing’ and ‘with physical contact’. ‘Other patients’ for these options referring to patients who did not have ‘confirmed or suspected COVID-19’. Midpoints were then added together across the three questions and groups were then categorised as before (0,1-5,6-20,21-50,51+). For the ‘51+’ option where there was no midpoint, ‘51’ was used for the purposes of response summation. We compared this variable with a variable that quantified total patient contact per week (regardless of COVID-19 status). Associations with mental health were quantitatively similar. |
| **Current working hours per week** | Continuous variable. Participants were asked: ‘At present, how many hours do you work in a typical week?’ |
| **Currently working at night** | Binary variable. Participants were asked how often they worked night shifts, with the following scale used for responses: Not applicable; Never; Less than once a month; Once a month or more, but not every week; Once a week or more, but not every shift; I always work nights. Participants were coded as ‘No’ if they selected ‘Not applicable’ or ‘Never’, and ‘Yes’ otherwise. |
| **Access to appropriate PPE** | Binary variable. Participants were asked to indicate how frequently they had access to PPE with responses: ‘Not at all’, ‘Rarely’, ‘Some of the time’, ‘Yes, most of the time’, ‘Yes, all of the time’, ‘Not applicable’. This was collapsed into the categorical variable ‘Rarely or not at all’, “Some or most of the time”, “Not applicable or all the time”. |

**Supplementary Figure 1.** Flowchart describing participant recruitment, consent, and derivation of analysis sample.

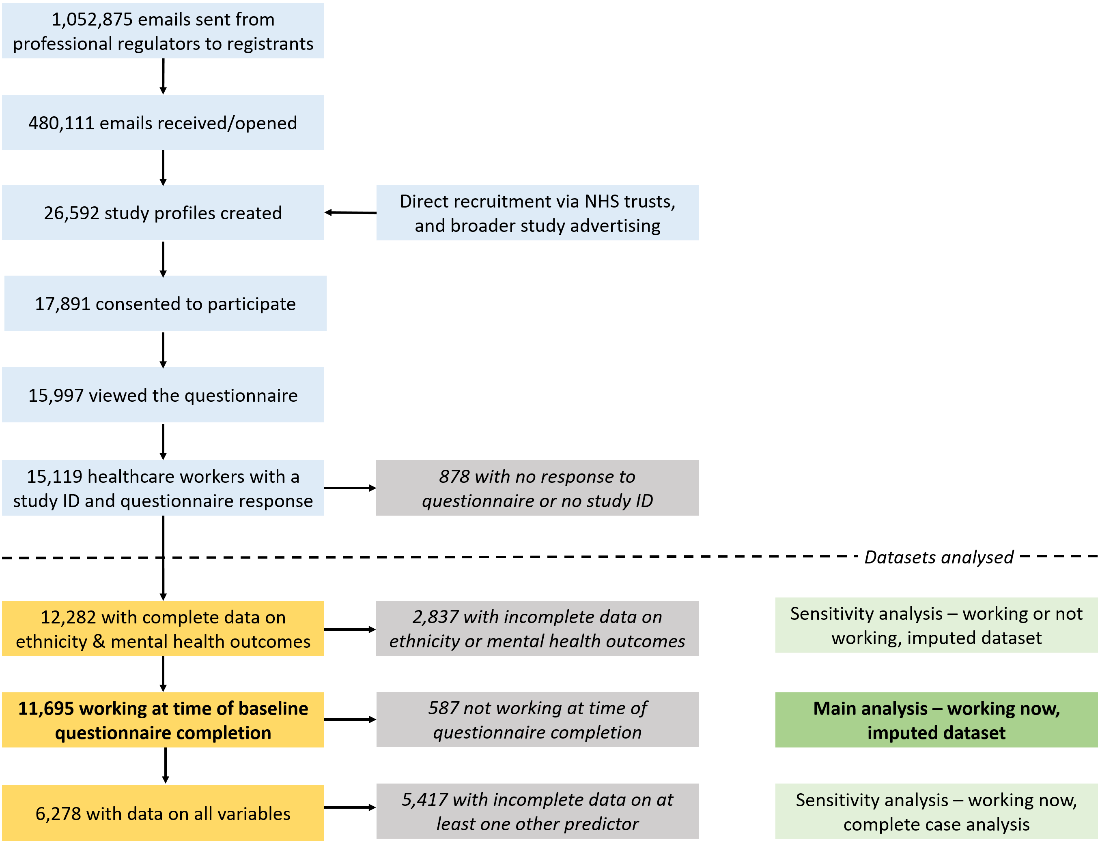

**Supplementary Table 1.** Sensitivity analyses for models shown in Table 2, using non-imputed data (but complete for confounders, max N=11,207).

| **Variable** | **N** | **Anxiety/depression symptoms** | | | | | | **PTSD symptoms** | | | | | |
| --- | --- | --- | --- | --- | --- | --- | --- | --- | --- | --- | --- | --- | --- |
|  |  | **Univariable** | | | **Adjusted for age, sex, ethnicity, job role** | | | **Univariable** | | | **Adjusted for age, sex, ethnicity, job role** | | |
|  |  | **OR** | **95%CI** | **P** | **OR** | **95%CI** | **P** | **OR** | **95%CI** | **P** | **OR** | **95%CI** | **P** |
| **Ethnicity**  White  Asian  Black  Mixed  Other | 11207 | ref  0.85  0.72  1  1.12 | 0.76,0.96  0.57,0.92  0.81,1.25  0.83,1.53 | 0.007  0.008  0.972  0.452 | ref  0.95  0.78  0.98  1.38 | 0.83,1.08  0.61,1.00  0.79,1.23  1.00,1.90 | 0.418  0.054  0.883  0.050 | ref  1.19  1.12  1.07  1.27 | 1.04,1.36  0.86,1.46  0.82,1.40  0.89,1.83 | 0.010  0.411  0.624  0.192 | ref  1.53  1.32  1.17  1.75 | 1.32,1.78  1.01,1.73  0.89,1.53  1.20,2.54 | 1.38E-08  0.044  0.266  0.004 |
| **Sex**  Male  Female | 11207 | ref  1.58 | 1.42,1.77 | <1E-10 | ref  1.29 | 1.15,1.46 | 1.94E-05 | ref  1.64 | 1.43,1.89 | <1E-10 | ref  1.41 | 1.21,1.63 | 5.62E-06 |
| **Age (per decade)** | 11207 | 0.69 | 0.66,0.72 | <1E-10 | 0.68 | 0.65,0.71 | <1E-10 | 0.79 | 0.76,0.83 | <1E-10 | 0.79 | 0.75,0.83 | <1E-10 |
| **Job**  Doctors and medical support  Nurses, NAs, Midwives  Allied Health Professionals and Pharmacists  Dental  Admin/estates/other | 11207 | ref  1.86  1.47  1.67  1.46 | 1.63,2.13  1.30,1.66  1.37,2.04  1.18,1.80 | <1E-10  4.25E-10  3.18E-07  0.001 | ref  1.9  1.31  1.49  1.36 | 1.64,2.21  1.15,1.50  1.21,1.82  1.09,1.69 | <1E-10  6.02E-05  1.55E-04  0.007 | ref  2.18  1.45  1.81  1.5 | 1.85,2.58  1.25,1.69  1.43,2.30  1.15,1.94 | <1E-10  1.78E-06  1.17E-06  0.003 | ref  2.55  1.54  1.83  1.67 | 2.12,3.06  1.30,1.81  1.43,2.34  1.27,2.19 | <1E-10  4.29E-07  1.62E-06  2.29E-04 |
| **Migration Status**  Not born in UK  Born in UK | 11181 | ref  1.11 | 1.00,1.23 | 0.047 | ref  0.96 | 0.85,1.08 | 0.526 | ref  0.94 | 0.83,1.06 | 0.301 | ref  0.94 | 0.81,1.08 | 0.356 |
| **Index of multiple deprivation** | 9923 | 0.93 | 0.92,0.95 | <1E-10 | 0.96 | 0.94,0.97 | 2.24E-06 | 0.92 | 0.90,0.94 | <1E-10 | 0.94 | 0.92,0.96 | 2.41E-08 |
| **Household size** | 11197 | 0.93 | 0.90,0.96 | 2.37E-05 | 0.93 | 0.90,0.96 | 5.03E-05 | 0.97 | 0.93,1.01 | 0.098 | 0.96 | 0.92,1.00 | 0.033 |
| **Religiosity**  Not at all important  Fairly important  Very important  Extremely important | 10963 | ref  0.87  0.93  0.81 | 0.78,0.98  0.80,1.08  0.69,0.94 | 0.019  0.353  0.006 | ref  0.98  1.12  0.91 | 0.87,1.11  0.95,1.32  0.77,1.07 | 0.751  0.191  0.262 | ref  1.19  1.39  1.19 | 1.04,1.36  1.17,1.66  1.00,1.43 | 0.013  2.53E-04  0.050 | ref  1.22  1.46  1.16 | 1.05,1.40  1.21,1.76  0.96,1.41 | 0.007  9.45E-05  0.125 |
| **Living with children**  Does not live with children  Lives with children | 9923 | ref  0.93 | 0.84,1.02 | 0.126 | ref  0.95 | 0.86,1.04 | 0.263 | ref  1.02 | 0.91,1.15 | 0.714 | ref  1.03 | 0.91,1.15 | 0.666 |
| **Living with adults 65+**  Does not live with adults 65+  Lives with adults 65+ | 9923 | ref  0.63 | 0.52,0.76 | 1.16E-06 | ref  0.85 | 0.70,1.03 | 0.104 | ref  0.87 | 0.71,1.08 | 0.212 | ref  1 | 0.81,1.25 | 0.968 |
| **Bereavement due to COVID-19**  Does not know someone who died  Knows someone who died | 11082 | ref  1.18 | 1.08,1.29 | 2.12E-04 | ref  1.3 | 1.18,1.42 | 4.71E-08 | ref  1.48 | 1.33,1.65 | <1E-10 | ref  1.51 | 1.35,1.69 | <1E-10 |
| **Long-term conditions** | 10771 | 1.17 | 1.07,1.27 | 0.001 | 1.31 | 1.19,1.43 | 8.11E-09 | 1.27 | 1.15,1.41 | 3.16E-06 | 1.36 | 1.23,1.51 | 7.07E-09 |
| **SARS-CoV-2 infection**  No SARS-CoV-2 infection  SARS-CoV-2 infection | 11150 | ref  1.06 | 0.95,1.18 | 0.277 | ref  0.97 | 0.87,1.08 | 0.560 | ref  1.07 | 0.94,1.21 | 0.323 | ref  1 | 0.87,1.14 | 0.957 |
| **Alcohol frequency**  Never  Monthly or less  2-4 times per month  2-3 times per week  4+ times per week | 11159 | ref  1.1  0.95  0.76  0.93 | 0.95,1.27  0.82,1.10  0.66,0.88  0.78,1.10 | 0.189  0.478  2.24E-04  0.401 | ref  0.96  0.81  0.77  1.14 | 0.83,1.11  0.69,0.94  0.65,0.90  0.94,1.37 | 0.572  0.007  0.001  0.186 | ref  1.03  0.8  0.62  0.79 | 0.88,1.22  0.68,0.95  0.52,0.74  0.64,0.98 | 0.693  0.011  1.38E-07  0.031 | ref  0.99  0.81  0.72  1.04 | 0.84,1.18  0.67,0.97  0.59,0.86  0.83,1.30 | 0.949  0.020  0.001  0.732 |
| **Smoking status**  Never-smoker  Ex-smoker  Current smoker | 11102 | ref  1.2  1.95 | 1.08,1.33  1.63,2.34 | 0.001  <1E-10 | ref  1.27  1.77 | 1.13,1.42  1.46,2.14 | 3.35E-05  3.58E-09 | ref  1.25  1.75 | 1.10,1.41  1.41,2.18 | 0.001  3.05E-07 | ref  1.31  1.62 | 1.15,1.50  1.30,2.03 | 6.93E-05  1.78E-05 |
| **Physical Activity Index (PAI)**  Inactive  Moderately inactive  Moderately active  Active | 10617 | ref  0.95  0.78  0.69 | 0.83,1.08  0.68,0.88  0.61,0.79 | 0.438  1.12E-04  2.66E-08 | ref  0.85  0.7  0.59 | 0.74,0.97  0.61,0.80  0.51,0.67 | 0.017  1.27E-07  <1E-10 | ref  1.05  0.86  0.82 | 0.89,1.23  0.73,1.00  0.70,0.96 | 0.587  0.055  0.015 | ref  0.94  0.82  0.77 | 0.80,1.11  0.69,0.96  0.65,0.90 | 0.466  0.014  0.002 |
| **Discrimination at work**  Not discriminated against at work  Discriminated against at work | 10404 | ref  2.25 | 2.04,2.47 | <1E-10 | ref  2.2 | 1.99,2.43 | <1E-10 | ref  2.85 | 2.54,3.19 | <1E-10 | ref  2.72 | 2.42,3.07 | <1E-10 |
| **Redeployment**  Not redeployed  Redeployed  Not working | 11155 | ref  1.22  1.1 | 1.09,1.37  0.96,1.26 | 0.001  0.162 | ref  1.13  0.95 | 1.01,1.27  0.82,1.10 | 0.039  0.487 | ref  1.2  1.2 | 1.05,1.38  1.03,1.41 | 0.010  0.024 | ref  1.14  1.09 | 0.99,1.31  0.92,1.29 | 0.067  0.332 |
| **Job areas**  Not working in area specified  Ambulance  Community clinical  Community non-clinical  Emergency department  Intensive care  Inpatient setting  Outpatient  Public / communal setting  Mobile across areas  Prison  Other clinical setting  Nursing or care home  Psychiatric hospital / inpatient unit  At home  University | 10946  10945  10947  10945  10942  10940  10939  10948  10947  10947  10939  10946  10947  10943  10946 | ref  1.22  0.86  1.24  1.27  1.4  1.16  0.86  1.49  1.04  1  1.05  0.88  1.36  1.06  1.08 | 0.97,1.52  0.77,0.95  1.04,1.49  1.08,1.48  1.19,1.66  1.05,1.28  0.76,0.96  1.15,1.94  0.81,1.34  0.55,1.83  0.93,1.19  0.65,1.19  1.05,1.77  0.94,1.19  0.81,1.44 | 0.083  0.003  0.017  0.003  7.40E-05  0.004  0.007  0.003  0.763  0.989  0.453  0.403  0.020  0.354  0.593 | ref  1.16  0.88  1.21  1.18  1.19  1.08  0.92  1.44  1.01  0.97  1.11  0.85  1.29  1.13  1.14 | 0.91,1.47  0.79,0.98  1.01,1.46  1.00,1.40  1.00,1.42  0.97,1.21  0.82,1.04  1.09,1.88  0.78,1.30  0.53,1.79  0.98,1.26  0.62,1.16  0.99,1.69  1.00,1.28  0.85,1.53 | 0.222  0.024  0.038  0.047  0.048  0.153  0.186  0.009  0.964  0.931  0.114  0.308  0.062  0.060  0.385 | ref  1.03  0.83  1.23  1.15  1.38  1.26  0.94  1.76  1.25  1.09  1.3  1.13  1.22  0.9  0.83 | 0.77,1.36  0.73,0.94  0.99,1.52  0.95,1.39  1.13,1.68  1.12,1.43  0.82,1.08  1.31,2.36  0.94,1.68  0.53,2.21  1.13,1.51  0.80,1.60  0.89,1.69  0.78,1.05  0.57,1.21 | 0.864  0.004  0.059  0.165  0.002  1.65E-04  0.413  1.64E-04  0.125  0.821  3.15E-04  0.488  0.217  0.185  0.336 | ref  1.18  0.85  1.21  1.16  1.27  1.27  1.05  1.86  1.3  1.11  1.37  1.05  1.17  0.96  0.89 | 0.88,1.59  0.74,0.97  0.98,1.51  0.95,1.42  1.04,1.56  1.12,1.45  0.91,1.21  1.38,2.52  0.97,1.75  0.54,2.28  1.18,1.59  0.74,1.49  0.85,1.62  0.82,1.12  0.60,1.30 | 0.269  0.014  0.083  0.152  0.022  2.84E-04  0.486  5.48E-05  0.077  0.775  2.86E-05  0.782  0.343  0.593  0.536 |
| **Secure about raising concerns related to clinical practice**  secure about raising concerns  not secure in raising concerns | 10868 | ref  2.13 | 1.89,2.38 | <1E-10 | ref  2.08 | 1.85,2.33 | <1E-10 | ref  2.17 | 1.92,2.5 | <1E-10 | ref  2.04 | 1.79,2.33 | <1E-10 |
| **Trusts in organisation to address concerns related to clinical practice**  trusts organisation  does not trust organisation | 10938 | ref  1.89 | 1.72,2.08 | <1E-10 | ref  1.89 | 1.72,2.08 | <1E-10 | ref  1.92 | 1.72,2.17 | <1E-10 | ref  1.89 | 1.60,2.13 | <1E-10 |
| **Non-COVID-19 patient contact**  0  1-5  6-20  21-50  51+ | 10959 | ref  1.06  0.96  0.97  0.99 | 0.85,1.33  0.82,1.12  0.84,1.12  0.85,1.14 | 0.590  0.587  0.673  0.844 | ref  1  0.89  0.89  0.96 | 0.79,1.25  0.76,1.05  0.76,1.04  0.82,1.12 | 0.970  0.162  0.158  0.593 | ref  0.77  0.78  0.94  1 | 0.58,1.02  0.65,0.94  0.79,1.12  0.84,1.19 | 0.066  0.010  0.496  0.965 | ref  0.72  0.74  0.9  1 | 0.54,0.96  0.61,0.90  0.75,1.09  0.83,1.20 | 0.023  0.003  0.283  0.984 |
| **COVID-19 patient contact**  0  1-5  6-20  21-50  51+ | 10998 | ref  1.15  1.16  1.45  2.15 | 0.96,1.36  1.03,1.31  1.23,1.70  1.71,2.70 | 0.120  0.011  5.35E-06  <1E-10 | ref  1.09  1.12  1.29  1.77 | 0.92,1.31  0.99,1.27  1.09,1.53  1.39,2.25 | 0.318  0.075  0.003  2.86E-06 | ref  1.39  1.28  1.56  2.38 | 1.14,1.70  1.11,1.47  1.29,1.89  1.84,3.08 | 0.001  0.001  3.88E-06  <1E-10 | ref  1.38  1.3  1.52  2.14 | 1.12,1.69  1.12,1.51  1.25,1.85  1.64,2.79 | 0.002  0.001  3.02E-05  2.67E-08 |
| **Working hours** | 11087 | 1.01 | 1.01,1.02 | 8.95E-09 | 1.01 | 1.01,1.02 | 1.13E-05 | 1.01 | 1.00,1.01 | 4.35E-04 | 1.01 | 1.01,1.02 | 2.02E-04 |
| **Working nights**  No  Yes | 10816 | ref  1.26 | 1.14,1.39 | 8.39E-06 | ref  1.16 | 1.04,1.30 | 0.008 | ref  1.26 | 1.11,1.42 | 2.28E-04 | ref  1.24 | 1.09,1.42 | 0.001 |
| **Access to PPE**  not applicable or all the time  some or most of the time  Rarely or not at all | 11181 | ref  1.67  1.12 | 1.48,1.88  0.67,1.85 | <1E-10  0.668 | ref  1.57  1.33 | 1.39,1.77  0.80,2.23 | <1E-10  0.275 | ref  1.81  0.81 | 1.58,2.08  0.41,1.62 | <1E-10  0.552 | ref  1.75  0.87 | 1.52,2.02  0.43,1.76 | <1E-10  0.704 |

**Supplementary Table 2.** Adjusted results from Table 2 presented alongside results additionally adjusted for month of consent (a proxy for completion date) for the imputed dataset of 11,695 individuals working now.

| **Variable** | **Anxiety/Depression** | | | | **PTSD** | | | |
| --- | --- | --- | --- | --- | --- | --- | --- | --- |
|  | **Adjusted**  **(Age, Sex, Ethnicity, Job role)** | | **Adjusted**  **(Age, Sex, Ethnicity, Job role, month consented)** | | **Adjusted**  **(Age, Sex, Ethnicity, Job role)** | | **Adjusted**  **(Age, Sex, Ethnicity, Job role, month consented)** | |
|  | **OR (95% CI)** | **P** | **OR (95% CI)** | **P** | **OR (95% CI)** | **P** | **OR (95% CI)** | **P** |
| **Ethnicity**  White  Asian  Black  Mixed  Other | ref  0.94 (0.83,1.07)  0.82 (0.65,1.04)  0.95 (0.76,1.18)  1.33 (0.98,1.80) | 0.351  0.095  0.618  0.070 | ref  0.94 (0.83,1.07)  0.81 (0.64,1.03)  0.95 (0.76,1.19)  1.33 (0.98,1.81) | 0.356  0.080  0.657  0.066 | ref  1.55 (1.34,1.78)  1.32 (1.01,1.71)  1.11 (0.85,1.46)  1.83 (1.29,2.60) | 2.13E-09  0.039  0.431  0.001 | ref  1.55 (1.34,1.79)  1.31 (1.01,1.70)  1.12 (0.86,1.46)  1.85 (1.30,2.63) | 1.7E-09  0.046  0.411  0.001 |
| **Sex**  Male  Female | ref  1.3 (1.16,1.46) | 8.6E-06 | ref  1.3 (1.16,1.46) | 8.8E-06 | ref  1.41 (1.22,1.63) | 3.2E-06 | ref  1.41 (1.22,1.63) | 3.4E-06 |
| **Age (per decade)** | 0.68 (0.66,0.71) | <1E-10 | 0.68 (0.66,0.71) | <1E-10 | 0.79 (0.75,0.83) | <1E-10 | 0.79 (0.76,0.83) | <1E-10 |
| **Job**  Doctors and medical support  Nurses, NAs, Midwives  Allied Health Professionals and Pharmacists  Dental  Admin/estates/other | ref  1.87 (1.61,2.18)  1.31 (1.14,1.49)  1.46 (1.20,1.79)  1.36 (1.10,1.70) | <1E-10  7.6E-05  0.0002  0.005 | ref  1.93 (1.65,2.25)  1.31 (1.14,1.49)  1.44 (1.17,1.76)  1.3 (1.04,1.63) | <1E-10  8.4E-05  0.0004  0.023 | ref  2.51 (2.09,3.01)  1.52 (1.29,1.79)  1.78 (1.40,2.27)  1.7 (1.30,2.21) | 0  5.9E-07  3.5E-06  0.0001 | ref  2.52 (2.09,3.03)  1.5 (1.27,1.77)  1.78 (1.39,2.27)  1.57 (1.19,2.07) | 3.0E-22  1.6E-06  4.3E-06  0.001 |
| **Migration Status**  Not born in UK  Born in UK | ref  0.96 (0.85,1.08) | 0.482 | ref  0.96 (0.85,1.08) | 0.484 | ref  0.9 (0.78,1.03) | 0.133 | ref  0.9 (0.78,1.03) | 0.134 |
| **Index of multiple deprivation** | 0.96 (0.94,0.98) | 3.8E-05 | 0.96 (0.94,0.98) | 3.5E-05 | 0.94 (0.92,0.96) | 5.3E-07 | 0.94 (0.92,0.96) | 4.7E-07 |
| **Household size** | 0.93 (0.90,0.96) | 2.1E-05 | 0.93 (0.90,0.96) | 1.4E-05 | 0.95 (0.91,0.99) | 0.015 | 0.95 (0.91,0.99) | 0.013 |
| **Religiosity**  Not at all important  Fairly important  Very important  Extremely important | ref  0.99 (0.88,1.11)  1.1 (0.94,1.30)  0.91 (0.77,1.07) | 0.872  0.229  0.246 | ref  0.99 (0.88,1.11)  1.1 (0.94,1.30)  0.91 (0.77,1.07) | 0.887  0.233  0.247 | ref  1.21 (1.05,1.39)  1.39 (1.16,1.67)  1.2 (0.99,1.44) | 0.007  0.0003  0.061 | ref  1.21 (1.05,1.39)  1.39 (1.16,1.67)  1.2 (0.99,1.44) | 0.007  0.0004  0.061 |
| **Living with children**  Does not live with children  Lives with children | ref  0.89 (0.81,0.98) | 0.014 | ref  0.89 (0.81,0.98) | 0.013 | ref  0.97 (0.87,1.09) | 0.646 | ref  0.97 (0.87,1.09) | 0.625 |
| **Living with adults 65+**  Does not live with adults 65+  Live with adults 65+ | ref  0.87 (0.73,1.05) | 0.153 | ref  0.88 (0.73,1.05) | 0.157 | ref  0.98 (0.79,1.22) | 0.847 | ref  0.98 (0.79,1.22) | 0.869 |
| **Bereavement due to COVID-19**  Does not know someone who died  Knows someone who died | ref  1.29 (1.18,1.42) | 2.6E-08 | ref  1.28 (1.17,1.41) | 8.3E-08 | ref  1.49 (1.34,1.67) | <1E-10 | ref  1.48 (1.33,1.65) | <1E-10 |
| **Long-term conditions** | 1.31 (1.20,1.42) | 2.2E-09 | 1.31 (1.20,1.43) | 1.7E-09 | 1.34 (1.21,1.48) | 2.6E-08 | 1.34 (1.21,1.49) | 2.6E-08 |
| **SARS-CoV-2 infection**  No SARS-CoV-2 infection  SARS-CoV-2 infection | ref  0.97 (0.87,1.07) | 0.518 | ref  0.96 (0.87,1.07) | 0.501 | ref  0.99 (0.87,1.12) | 0.821 | ref  0.98 (0.86,1.12) | 0.790 |
| **Alcohol frequency**  Never  Monthly or less  2-4 times per month  2-3 times per week  4+ times per week | ref  0.95 (0.82,1.10)  0.81 (0.69,0.94)  0.76 (0.65,0.89)  1.12 (0.93,1.35) | 0.489  0.005  0.001  0.235 | ref  0.95 (0.82,1.10)  0.8 (0.69,0.93)  0.76 (0.65,0.89)  1.12 (0.93,1.35) | 0.490  0.004  0.001  0.221 | ref  0.96 (0.81,1.13)  0.78 (0.65,0.93)  0.68 (0.57,0.82)  1.01 (0.81,1.26) | 0.601  0.005  4.5E-05  0.915 | ref  0.96 (0.81,1.13)  0.78 (0.65,0.93)  0.68 (0.57,0.82)  1.02 (0.81,1.27) | 0.608  0.005  4.8E-05  0.894 |
| **Smoking status**  Never-smoker  Ex-smoker  Current smoker | ref  1.24 (1.11,1.38)  1.76 (1.46,2.12) | 0.0001  2.3E-09 | ref  1.24 (1.11,1.38)  1.76 (1.46,2.12) | 0.0001  2.4E-09 | ref  1.29 (1.13,1.47)  1.66 (1.34,2.06) | 0.0001  4.3E-06 | ref  1.29 (1.13,1.47)  1.66 (1.34,2.06) | 0.0001  4.1E-06 |
| **Physical Activity Index (PAI)**  Inactive  Moderately inactive  Moderately active  Active | ref  0.86 (0.75,0.98)  0.71 (0.62,0.81)  0.6 (0.53,0.69) | 0.024  1.9E-07  <1E-10 | ref  0.86 (0.75,0.98)  0.71 (0.63,0.81)  0.6 (0.53,0.69) | 0.024  2.7E-07  <1E-10 | ref  0.97 (0.82,1.14)  0.83 (0.71,0.97)  0.8 (0.68,0.94) | 0.683  0.018  0.006 | ref  0.97 (0.82,1.14)  0.83 (0.71,0.97)  0.8 (0.68,0.94) | 0.690  0.020  0.007 |
| **Discrimination at work**  Not discriminated against at work  Discriminated against at work | ref  2.12 (1.92,2.33) | <1E-10 | ref  2.13 (1.93,2.35) | <1E-10 | ref  2.64 (2.35,2.95) | <1E-10 | ref  2.65 (2.36,2.97) | <1E-10 |
| **Redeployment**  Not redeployed  Redeployed  Not working | ref  1.15 (1.03,1.29)  0.95 (0.83,1.10) | 0.015  0.518 | ref  1.15 (1.03,1.29)  0.96 (0.83,1.10) | 0.015  0.553 | ref  1.17 (1.02,1.34)  1.09 (0.92,1.28) | 0.028  0.331 | ref  1.17 (1.02,1.34)  1.09 (0.93,1.29) | 0.029  0.297 |
| **Job areas**  Not working in area specified  Ambulance  Community clinical  Community non-clinical  Emergency department  Intensive care  Inpatient setting  Outpatient  Public / communal setting  Mobile across areas  Prison  Other clinical setting  Nursing or care home  Psychiatric hospital / inpatient unit  At home  University | ref  1.13 (0.90,1.42)  0.88 (0.80,0.98)  1.19 (0.99,1.43)  1.15 (0.97,1.35)  1.19 (1.00,1.41)  1.05 (0.95,1.17)  0.94 (0.83,1.05)  1.36 (1.04,1.78)  1 (0.78,1.30)  1.03 (0.57,1.87)  1.09 (0.96,1.24)  0.9 (0.67,1.22)  1.26 (0.96,1.64)  1.14 (1.01,1.28)  1.13 (0.84,1.50) | 0.291  0.019  0.061  0.104  0.049  0.347  0.268  0.024  0.974  0.931  0.164  0.500  0.090  0.041  0.418 | ref  1.14 (0.90,1.43)  0.89 (0.80,0.98)  1.2 (1.00,1.44)  1.15 (0.98,1.35)  1.19 (1.00,1.41)  1.05 (0.95,1.17)  0.93 (0.83,1.05)  1.35 (1.04,1.77)  1.01 (0.78,1.30)  1.01 (0.56,1.84)  1.09 (0.96,1.24)  0.91 (0.68,1.23)  1.24 (0.95,1.62)  1.13 (1.00,1.28)  1.13 (0.85,1.50) | 0.277  0.023  0.054  0.096  0.047  0.342  0.246  0.026  0.939  0.972  0.169  0.548  0.108  0.048  0.414 | ref  1.18 (0.89,1.57)  0.84 (0.74,0.95)  1.15 (0.93,1.43)  1.13 (0.93,1.38)  1.28 (1.04,1.56)  1.21 (1.07,1.38)  1.04 (0.90,1.19)  1.76 (1.31,2.37)  1.3 (0.97,1.73)  1.22 (0.61,2.45)  1.37 (1.19,1.59)  1.14 (0.82,1.58)  1.13 (0.82,1.56)  0.95 (0.81,1.10)  0.91 (0.62,1.32) | 0.250  0.006  0.200  0.209  0.017  0.003  0.626  0.0001  0.077  0.570  1.9E-05  0.445  0.471  0.483  0.605 | ref  1.18 (0.89,1.57)  0.84 (0.74,0.95)  1.16 (0.93,1.44)  1.14 (0.93,1.38)  1.28 (1.05,1.56)  1.21 (1.07,1.38)  1.03 (0.90,1.18)  1.75 (1.30,2.35)  1.3 (0.98,1.74)  1.21 (0.60,2.42)  1.37 (1.19,1.59)  1.15 (0.83,1.60)  1.11 (0.80,1.53)  0.94 (0.81,1.10)  0.9 (0.62,1.32) | 0.246  0.007  0.181  0.198  0.016  0.003  0.676  0.0002  0.072  0.598  2.2E-05  0.401  0.539  0.441  0.601 |
| **Secure about raising concerns related to clinical practice**  secure about raising concerns  not secure in raising concerns | ref  2.04 (1.81,2.27) | <1E-10 | ref  2.04 (1.81,2.27) | <1E-10 | ref  2.08 (1.81,2.38) | <1E-10 | ref  2.08 (1.85,2.38) | <1E-10 |
| **Trusts in organisation to address concerns related to clinical practice**  trusts organisation  does not trust organisation | ref  1.89 (1.72,2.08) | <1E-10 | ref  1.89 (1.72,2.08) | <1E-10 | ref  1.89 (1.60,2.08) | <1E-10 | ref  1.89 (1.69,2.13) | <1E-10 |
| **Non-COVID-19 patient contact**  0  1-5  6-20  21-50  51+ | ref  1 (0.80,1.25)  0.88 (0.75,1.03)  0.89 (0.76,1.04)  0.95 (0.81,1.11) | 0.100  0.099  0.132  0.505 | ref  0.99 (0.79,1.24)  0.87 (0.74,1.02)  0.89 (0.77,1.04)  0.95 (0.82,1.11) | 0.931  0.094  0.146  0.544 | ref  0.71 (0.54,0.95)  0.74 (0.61,0.89)  0.91 (0.76,1.09)  0.99 (0.83,1.19) | 0.021  0.002  0.297  0.956 | ref  0.7 (0.53,0.94)  0.73 (0.61,0.89)  0.91 (0.76,1.09)  1 (0.84,1.20) | 0.016  0.001  0.311  1.000 |
| **COVID-19 patient contact**  0  1-5  6-20  21-50  51+ | ref  1.09 (0.92,1.30)  1.13 (1.00,1.28)  1.3 (1.11,1.53)  1.74 (1.38,2.19) | 0.332  0.048  0.001  2.5E-06 | ref  1.09 (0.91,1.29)  1.13 (1.00,1.28)  1.3 (1.10,1.53)  1.72 (1.36,2.16) | 0.348  0.046  0.002  4.5E-06 | ref  1.36 (1.11,1.67)  1.33 (1.16,1.54)  1.52 (1.26,1.85)  2.08 (1.61,2.70) | 0.003  9.0E-05  1.6E-05  2.6E-08 | ref  1.36 (1.11,1.66)  1.34 (1.16,1.55)  1.52 (1.26,1.85)  2.07 (1.60,2.68) | 0.003  8.1E-05  1.6E-05  3.7E-08 |
| **Working hours** | 1.01 (1.01,1.02) | 2.1E-05 | 1.01 (1.01,1.02) | 2.2E-05 | 1.01 (1.00,1.02) | 0.0003 | 1.01 (1.00,1.02) | 0.0003 |
| **Working nights**  No  Yes | ref  1.15 (1.03,1.28) | 0.012 | ref  1.16 (1.04,1.29) | 0.009 | ref  1.25 (1.09,1.42) | 0.001 | ref  1.26 (1.10,1.43) | 0.001 |
| **Access to PPE**  not applicable or all the time  some or most of the time  Rarely or not at all | ref  1.56 (1.39,1.76)  1.27 (0.76,2.12) | <1E-10  0.360 | ref  1.57 (1.39,1.77)  1.27 (0.76,2.13) | <1E-10  0.356 | ref  1.78 (1.55,2.04)  0.82 (0.41,1.66) | <1E-10  0.586 | ref  1.79 (1.56,2.05)  0.83 (0.41,1.67) | <1E-10  0.601 |
| **Date of consent**  December 2020  January 2021  February / March 2021 | - | - | ref  1.13 (1.02,1.26)  1.2 (1.06,1.36) | 0.024  0.003 | - | - | ref  1.06 (0.93,1.20)  1.2 (1.04,1.38) | 0.403  0.014 |

**Supplementary Table 3.** Associations between multiple non-work factors and symptoms of anxiety/depression and PTSD in 12,282 healthcare workers, regardless of working status at questionnaire completion.

| **Variable** | **Anxiety/Depression** | | | | **PTSD** | | | |
| --- | --- | --- | --- | --- | --- | --- | --- | --- |
|  | **Univariable** | | **Adjusted**  **(Age, Sex, Ethnicity, Job role)** | | **Univariable** | | **Adjusted**  **(Age, Sex, Ethnicity, Job role)** | |
|  | **OR (95% CI)** | **P** | **OR (95% CI)** | **P** | **OR (95% CI)** | **P** | **OR (95% CI)** | **P** |
| **Ethnicity**  White  Asian  Black  Mixed  Other | ref  0.86 (0.77,0.96)  0.73 (0.58,0.91)  1 (0.82,1.24)  1.11 (0.83,1.47) | 0.007  0.006  0.967  0.477 | ref  0.96 (0.85,1.08)  0.78 (0.62,0.99)  0.98 (0.79,1.21)  1.36 (1.01,1.82) | 0.486  0.038  0.852  0.044 | ref  1.19 (1.05,1.36)  1.15 (0.90,1.47)  1.14 (0.89,1.46)  1.39 (1.00,1.92) | 0.006  0.263  0.301  0.048 | ref  1.53 (1.33,1.76)  1.34 (1.04,1.72)  1.24 (0.96,1.59)  1.91 (1.36,2.68) | 2.3E-09  0.022  0.099  0.0002 |
| **Sex**  Male  Female | ref  1.59 (1.43,1.77) | <1E-10 | ref  1.28 (1.15,1.44) | 1.6E-05 | ref  1.67 (1.46,1.90) | <1E-10 | ref  1.43 (1.24,1.64) | 7.5E-07 |
| **Age (per decade)** | 0.7 (0.68,0.73) | <1E-10 | 0.69 (0.67,0.72) | <1E-10 | 0.8 (0.76,0.83) | <1E-10 | 0.8 (0.76,0.83) | <1E-10 |
| **Job**  Doctors and medical support  Nurses, NAs, Midwives  Allied Health Professionals and Pharmacists  Dental  Admin/estates/other | ref  1.88 (1.65,2.14)  1.46 (1.30,1.65)  1.65 (1.37,1.99)  1.44 (1.17,1.77) | <1E-10  1.9E-10  2.2E-07  0.001 | ref  1.92 (1.66,2.23)  1.32 (1.16,1.50)  1.47 (1.21,1.79)  1.36 (1.09,1.69) | <1E-10  2.4E-05  0.0001  0.006 | ref  2.16 (1.84,2.53)  1.46 (1.26,1.70)  1.78 (1.42,2.24)  1.49 (1.15,1.91) | <1E-10  4.7E-07  7.8E-07  0.002 | ref  2.51 (2.10,3.00)  1.55 (1.32,1.82)  1.8 (1.42,2.27)  1.66 (1.28,2.16) | <1E-10  7.8E-08  1.1E-06  0.0002 |
| **Migration Status**  Not born in UK  Born in UK | ref  1.09 (0.99,1.20) | 0.066 | ref  0.95 (0.85,1.07) | 0.400 | ref  0.91 (0.81,1.02) | 0.104 | ref  0.92 (0.80,1.05) | 0.197 |
| **Index of multiple deprivation** | 0.93 (0.92,0.95) | <1E-10 | 0.96 (0.94,0.97) | 2.7E-07 | 0.92 (0.90,0.94) | <1E-10 | 0.94 (0.92,0.96) | 8.3E-09 |
| **Household size** | 0.93 (0.90,0.96) | 9.8E-06 | 0.93 (0.90,0.96) | 4.6E-06 | 0.96 (0.93,1.00) | 0.066 | 0.95 (0.91,0.99) | 0.011 |
| **Religiosity**  Not at all important  Fairly important  Very important  Extremely important | ref  0.88 (0.79,0.98)  0.93 (0.81,1.08)  0.83 (0.72,0.96) | 0.022  0.359  0.014 | ref  0.98 (0.88,1.10)  1.11 (0.95,1.29)  0.94 (0.80,1.10) | 0.750  0.201  0.417 | ref  1.18 (1.04,1.34)  1.35 (1.14,1.59)  1.26 (1.07,1.48) | 0.012  0.001  0.006 | ref  1.2 (1.05,1.37)  1.39 (1.16,1.66)  1.22 (1.02,1.46) | 0.007  0.0003  0.027 |
| **Living with children**  Does not live with children  Lives with children | ref  0.88 (0.80,0.96) | 0.003 | ref  0.86 (0.79,0.94) | 0.001 | ref  0.98 (0.88,1.09) | 0.743 | ref  0.97 (0.87,1.08) | 0.546 |
| **Living with adults 65+**  Does not live with adults 65+  Lives with adults 65+ | ref  0.67 (0.56,0.79) | 4.0E-06 | ref  0.92 (0.76,1.10) | 0.336 | ref  0.87 (0.72,1.05) | 0.136 | ref  1.01 (0.83,1.23) | 0.905 |
| **Bereavement due to COVID-19**  Does not know someone who died  Knows someone who died | ref  1.22 (1.12,1.33) | 3.8E-06 | ref  1.33 (1.21,1.45) | 4.2E-10 | ref  1.51 (1.36,1.67) | <1E-10 | ref  1.52 (1.37,1.69) | <1E-10 |
| **Long-term conditions** | 1.18 (1.09,1.28) | 6.8E-05 | 1.32 (1.21,1.44) | 1.1E-10 | 1.29 (1.17,1.41) | 9.0E-08 | 1.38 (1.25,1.52) | <1E-10 |
| **SARS-CoV-2 infection**  No SARS-CoV-2 infection  SARS-CoV-2 infection | ref  1.08 (0.98,1.20) | 0.110 | ref  0.99 (0.89,1.10) | 0.863 | ref  1.06 (0.94,1.20) | 0.311 | ref  0.99 (0.88,1.12) | 0.890 |
| **Alcohol frequency**  Never  Monthly or less  2-4 times per month  2-3 times per week  4+ times per week | ref  1.08 (0.95,1.24)  0.93 (0.82,1.07)  0.75 (0.65,0.85)  0.88 (0.75,1.04) | 0.238  0.313  2.2E-05  0.138 | ref  0.95 (0.83,1.09)  0.81 (0.70,0.93)  0.76 (0.65,0.88)  1.09 (0.91,1.30) | 0.472  0.004  0.0003  0.349 | ref  1 (0.86,1.17)  0.76 (0.65,0.89)  0.59 (0.50,0.69)  0.75 (0.61,0.91) | 0.959  0.001  2.3E-10  0.003 | ref  0.97 (0.83,1.14)  0.77 (0.65,0.92)  0.68 (0.57,0.81)  0.98 (0.80,1.22) | 0.715  0.003  2.00E-05  0.884 |
| **Smoking status**  Never-smoker  Ex-smoker  Current smoker | ref  1.18 (1.07,1.30)  1.92 (1.61,2.28) | 0.001  <1E-10 | ref  1.25 (1.12,1.39)  1.74 (1.45,2.09) | 5.6E-05  2.1E-09 | ref  1.21 (1.07,1.36)  1.8 (1.47,2.21) | 0.002  1.2E-08 | ref  1.27 (1.12,1.45)  1.69 (1.37,2.08) | 0.0002  9.0E-07 |
| **Physical Activity Index (PAI)**  Inactive  Moderately inactive  Moderately active  Active | ref  0.96 (0.85,1.09)  0.79 (0.70,0.90)  0.71 (0.63,0.80) | 0.554  0.0002  6.8E-08 | ref  0.86 (0.76,0.98)  0.72 (0.63,0.81)  0.6 (0.53,0.69) | 0.025  2.4E-07  <1E-10 | ref  1.07 (0.92,1.25)  0.86 (0.74,1.00)  0.83 (0.71,0.96) | 0.392  0.056  0.013 | ref  0.97 (0.83,1.13)  0.83 (0.70,0.97)  0.78 (0.66,0.91) | 0.679  0.017  0.001 |

**Supplementary Table 4.** Results of serially adjusted analysis (Models 1-4), in 11,695 participants, as shown in Figure 1.

|  | **Anxiety/Depression** | | | | | | | | | |
| --- | --- | --- | --- | --- | --- | --- | --- | --- | --- | --- |
|  | **Univariable** | | **Model 1** | | **Model 2** | | **Model 3** | | **Model 4** | |
|  | **OR (95%CI)** | **P** | **OR (95%CI)** | **P** | **OR (95%CI)** | **P** | **OR (95%CI)** | **P** | **OR (95%CI)** | **P** |
| **Ethnicity**  White  Asian  Black  Mixed  Other | ref  0.85 (0.76,0.95)  0.76 (0.60,0.96) 0.97 (0.78,1.20) 1.09 (0.81,1.46) | 0.005  0.019  0.787  0.577 | ref  0.94 (0.83,1.07) 0.82 (0.65,1.04) 0.95 (0.76,1.18) 1.33 (0.98,1.80) | 0.351  0.095  0.618  0.070 | ref  0.9 (0.78,1.04) 0.77 (0.59,0.99) 0.92 (0.74,1.16) 1.24 (0.90,1.71) | 0.168  0.043  0.489  0.189 | ref  0.87 (0.75,1.01) 0.76 (0.59,0.98) 0.88 (0.70,1.10) 1.18 (0.85,1.63) | 0.068  0.036  0.273  0.316 | ref  0.77 (0.66,0.90) 0.65 (0.50,0.85) 0.81 (0.64,1.02) 1.02 (0.73,1.43) | 0.001  0.002  0.072  0.895 |
|  | **PTSD** | | | | | | | | | |
|  | **Univariable** | | **Model 1** | | **Model 2** | | **Model 3** | | **Model 4** | |
|  | **OR (95%CI)** | **P** | **OR (95%CI)** | **P** | **OR (95%CI)** | **P** | **OR (95%CI)** | **P** | **OR (95%CI)** | **P** |
| **Ethnicity**  White  Asian  Black  Mixed  Other | ref  1.21 (1.07,1.38)  1.13 (0.87,1.46)  1.03 (0.79,1.34)  1.34 (0.95,1.88) | 0.004  0.355  0.831  0.092 | ref  1.55 (1.34,1.78)  1.32 (1.01,1.71)  1.11 (0.85,1.46)  1.83 (1.29,2.60) | 2.1E-09  0.039  0.431  0.001 | ref  1.29 (1.10,1.52)  1.02 (0.77,1.36)  1.06 (0.81,1.39)  1.46 (1.01,2.11) | 0.002  0.876  0.674  0.045 | ref  1.27 (1.07,1.50)  1.02 (0.77,1.36)  1 (0.76,1.31)  1.41 (0.97,2.04) | 0.007  0.878  1.000  0.071 | ref  1.11 (0.93,1.33)  0.86 (0.64,1.15)  0.93 (0.70,1.23)  1.18 (0.80,1.73) | 0.236  0.307  0.594  0.399 |

**Supplementary Table 5.** Results of serially adjusted analysis (Models 1-4), in 6,278 participants with complete data for all variables.

|  | **Anxiety/Depression** | | | | | | | | | |
| --- | --- | --- | --- | --- | --- | --- | --- | --- | --- | --- |
|  | **Univariable** | | **Model 1** | | **Model 2** | | **Model 3** | | **Model 4** | |
|  | **OR (95%CI)** | **P** | **OR (95%CI)** | **P** | **OR (95%CI)** | **P** | **OR (95%CI)** | **P** | **OR (95%CI)** | **P** |
| **Ethnicity**  White  Asian  Black  Mixed  Other | ref  0.79 (0.68,0.93)  0.63 (0.45,0.88)  0.99 (0.75,1.32)  1.34 (0.91,1.99) | 0.004  0.008  0.967  0.139 | ref  0.9 (0.76,1.07)  0.69 (0.48,0.98)  0.96 (0.72,1.29)  1.77 (1.18,2.68) | 0.230  0.038  0.805  0.006 | ref  0.88 (0.72,1.07)  0.66 (0.46,0.97)  0.96 (0.72,1.28)  1.73 (1.13,2.67) | 0.209  0.034  0.778  0.013 | ref  0.86 (0.70,1.05)  0.66 (0.45,0.97)  0.9 (0.67,1.21)  1.67 (1.08,2.58) | 0.139  0.034  0.498  0.022 | ref  0.75 (0.61,0.93)  0.57 (0.39,0.85)  0.84 (0.62,1.13)  1.4 (0.89,2.21) | 0.009  0.005  0.249  0.142 |
|  | **PTSD** | | | | | | | | | |
|  | **Univariable** | | **Model 1** | | **Model 2** | | **Model 3** | | **Model 4** | |
|  | **OR (95%CI)** | **P** | **OR (95%CI)** | **P** | **OR (95%CI)** | **P** | **OR (95%CI)** | **P** | **OR (95%CI)** | **P** |
| **Ethnicity**  White  Asian  Black  Mixed  Other | ref  1.15 (0.96,1.38)  1.04 (0.71,1.52)  1.01 (0.71,1.44)  1.38 (0.85,2.22) | 0.135  0.838  0.955  0.191 | ref  1.5 (1.23,1.83)  1.26 (0.85,1.85)  1.1 (0.76,1.58)  2.01 (1.22,3.30) | 7.2E-05  0.245  0.611  0.006 | ref  1.25 (0.99,1.58)  0.97 (0.64,1.48)  1.05 (0.73,1.51)  1.6 (0.95,2.70) | 0.062  0.884  0.803  0.078 | ref  1.29 (1.01,1.64)  1 (0.65,1.53)  0.97 (0.67,1.40)  1.59 (0.94,2.70) | 0.041  0.995  0.871  0.083 | ref  1.11 (0.86,1.42)  0.84 (0.54,1.30)  0.91 (0.62,1.33)  1.31 (0.76,2.26) | 0.431  0.428  0.612  0.324 |
